# CLIN-SUMM: Incremental Longitudinal Summarization of Clinical Notes Enables Scalable Representation and Early Disease Prediction

**DOI:** 10.64898/2025.11.28.25341233

**Authors:** Valentina D’Souza, Danielle F. Pace, Alaleh Azhir, Arash Nargesi, Erik B. Holbrook, Wei He, Tristan Naumann, Samuel Friedman, Steven J. Atlas, Christopher D. Anderson, Judy Hung, Mahnaz Maddah

**Author notes:** Corresponding Author: Mahnaz Maddah, PhD, Director, Machine Learning for Health, Broad Institute of MIT and Harvard, 415 Main Street, Cambridge, MA 02142. Authors contributed equally to this project (Valentina D’Souza, Danielle F. Pace).

## Abstract

Electronic health records contain years of longitudinal clinical notes rich in evolving patient information, yet their volume, redundancy, and fragmentation limit clinical usability and scalable modeling. We present CLIN-SUMM (Clinical Longitudinal Insight from Notes using Summarization), a framework that restructures summarization as a longitudinal representation problem. Rather than collapsing histories into static summaries, CLIN-SUMM incrementally constructs structured, categorized, date-partitioned patient representations, summarizing only newly documented information at each encounter while preserving temporal fidelity without access to future data. This standardized representation can be computed once and reused across downstream tasks, thereby decoupling narrative processing from prediction. Across 12,356 Massachusetts General Hospital patients, CLIN-SUMM achieved 75% word reduction while maintaining high clinician-rated correctness and completeness. Using dementia as a case study, fine-tuning Clinical ModernBERT on CLIN-SUMM summaries yielded AUROC 0.84 for diagnosis and outperformed raw notes for 3-year risk prediction (AUROC 0.66 vs. 0.63). Combining summary-based predictions with structured EHR variables improved 3-year prediction to AUROC 0.69, reaching 0.74 among patients younger than 75 years, with longitudinal analyses demonstrating progressive risk separation years before a formal diagnosis. CLIN-SUMM summaries also enabled efficient extraction of longitudinal medication trajectories, complementing structured EHR data by recovering medication information documented in notes, with high dose agreement for matched records. CLIN-SUMM provides a scalable representation layer for clinical review and longitudinal machine learning, enhancing disease modeling, risk prediction, and other longitudinal reasoning tasks.

## Introduction

Electronic health records (EHRs) contain years of longitudinal clinical notes documenting the evolving trajectory of a patient’s medical history. These narratives capture symptoms, diagnoses, treatment decisions, responses to therapy, lifestyle factors, labs, imaging, and contextual clinical reasoning that are often incompletely represented in the structured codes typically used for machine learning ^1–3^. Clinical notes, therefore, are likely to remain narrative documents containing essential clinical information that can support a range of downstream tasks, including automated diagnosis, risk prediction, phenotyping, trajectory analysis, and clinical question answering.

Despite their richness, longitudinal notes remain difficult to use effectively at scale, whether by clinicians doing chart review or by machine learning models attempting to learn temporal patterns. The most obvious challenge is sheer volume: records of individual patients frequently accumulate hundreds to thousands of notes over many years. There is substantial redundancy due to use of note templates and copy-forward practices, with some estimating that over 50% of text is duplicated over encounters ^4^. The most critical clinical updates, such as new diagnoses, medication adjustments, or symptom progression, are often buried within thousands of lines of unstructured narrative text ^5^. Fragmentation compounds the problem: clinical observations are scattered across encounters and specialties, and individual notes rarely integrate longitudinal context into a coherent temporal account. Consequently, both clinicians and algorithms face substantial barriers when attempting to extract coherent longitudinal insight over unstructured narratives.

Although large language models (LLMs) aim to process ever-larger portions of the patient record, temporal reasoning in the longitudinal EHR remains a challenge. Kruse *et al.* ^6^ investigated high-level clinical summarization and future risk prediction from longitudinal structured and unstructured data in the MIMIC-III ICU dataset, finding that temporal reasoning over long histories remained suboptimal despite long context window, retrieval-augmented generation (RAG) and chain of thought (CoT) techniques. TIMER ^7^ fine-tunes a base LLM via supervised instruction tuning, using instruction-response pairs that explicitly focus on temporal patterns and are linked to corresponding timestamps. While this approach improved the LLM’s ability to answer questions requiring temporal reasoning, overall scores remained low.

We hypothesize that first addressing the large number of input tokens, low signal-to-noise ratios, redundancy and heterogeneous formatting of unstructured clinical notes will enable clinicians and models to use the record more accurately and efficiently. This is in the spirit of addressing signal dilution, the “lost in the middle” ^8^ effect and inference cost via redundancy removal and input text organization. For example, TRACE reduces “note bloat” by using EHR attribution metadata and frequency-based text flagging to remove templated and copy-forwarded text, maintaining information extraction and outcome prediction performance but leading to substantial projected cost savings ^9^.

However, most large language model (LLM) applications addressing clinical note summarization, whether extractive ^10^ or abstractive ^11^, typically operate only at the level of individual notes ^12^ or episodes (e.g., a hospitalization) ^13^, or collapse multi-visit histories into static summaries that obscure temporal evolution. Multi-modal approaches combining structured EHR data with clinical notes similarly operate only at the note or episode level ^14^, while methods that derive structured concepts from unstructured text lack the nuance contained within clinical narratives while remaining repetitive and unorganized ^15,16^.

The central challenge is therefore not simply summarization, but representation: how can we reliably transform large volumes of longitudinal clinical text into an organized, concise format that preserves the evolving clinical context, while remaining clinically navigable for chart review and computationally tractable for downstream modeling?

We introduce CLIN-SUMM (Clinical Longitudinal Insight from Notes using Summarization), a framework designed to generate structured, categorized, date-partitioned patient-level summaries through sequential summarization of multi-visit clinical notes (**Figure 1**). CLIN-SUMM does not simply “squash” a patient’s history into a paragraph, as in conventional summarization. At each encounter, CLIN-SUMM populates predefined clinical sections (e.g., “Diagnosis”, “Medications & Allergies”, “Vital signs or Lab/Imaging Findings” and more) with summaries of only the <u>new</u> information arising from that specific visit and appends date-stamped updates to explicitly capture changes over time.

**Figure 1:**
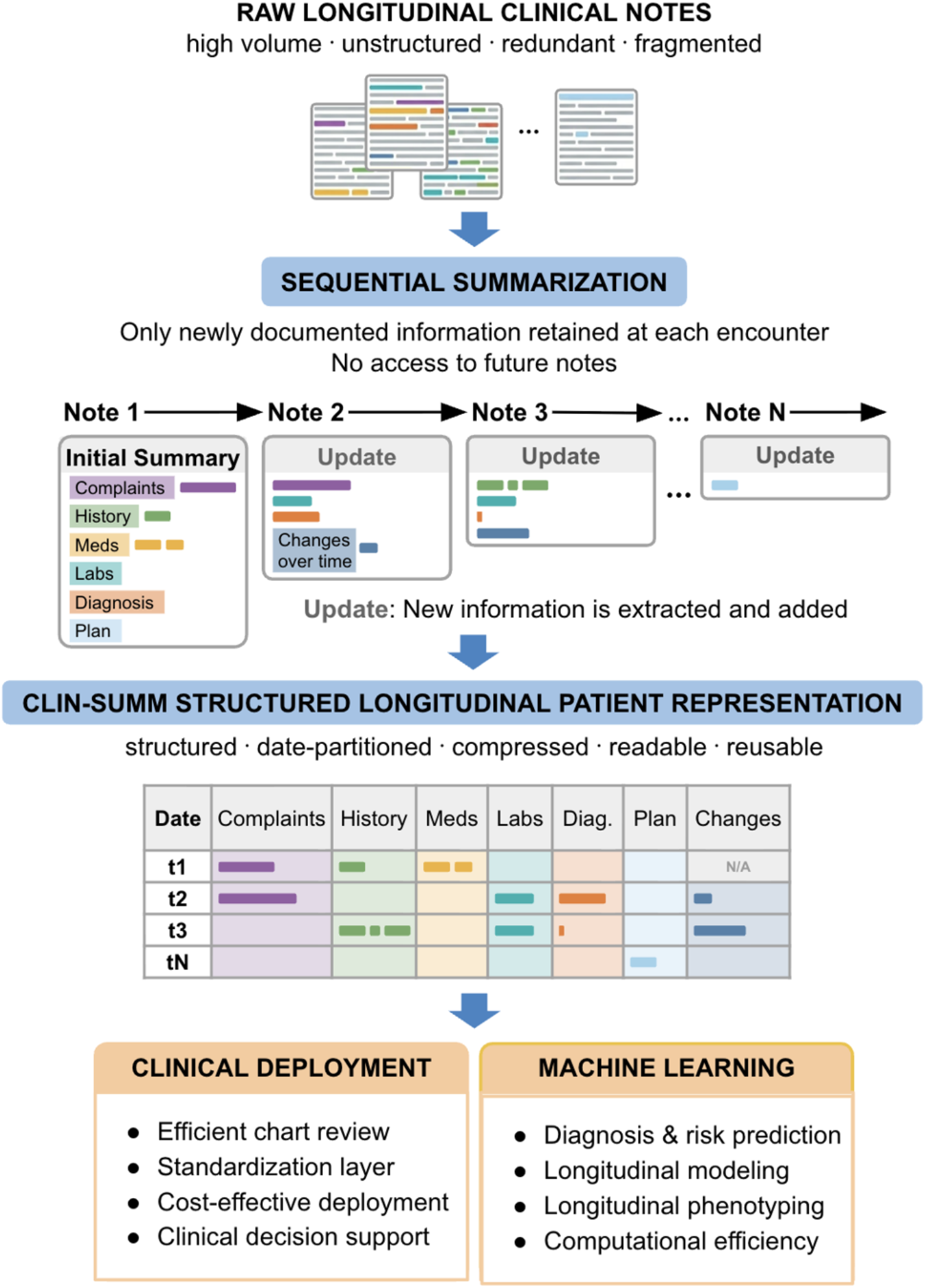
Overview of the CLIN-SUMM framework. The resulting patient-level data for each patient includes a summary of the first clinical note followed by summaries of the new updates described in subsequent notes, where each section captures how a patient’s clinical state evolves across encounters with corresponding dates. Patient visit notes are processed using LLM-powered summarization prompts, redundancy filtering, and sliding-window management to generate a longitudinal, date-partitioned clinical summary.

The resulting representation grows cumulatively as care continues, forming a longitudinal, structured summary of the patient’s health trajectory. In this sense, CLIN-SUMM encodes computationally how clinicians mentally update their understanding of a patient: new information refines and extends prior knowledge rather than rewriting history. Notes are processed strictly in chronological order. At every step the model updates the patient-level summary using only information available up to that time point, and the system never has access to “future” documentation when summarizing a given encounter. This preserves temporal fidelity and causal direction, aligning the representation with real-world clinical decision-making. Because CLIN-SUMM updates patient summaries after each encounter, a hospital system could feasibly generate and maintain longitudinal summaries as part of routine documentation in near real-time.

Conceptually, CLIN-SUMM functions as a representation learning layer between raw narrative text and downstream modeling tasks. Rather than training task-specific models directly on the original highly variable and repetitive clinical notes, or compressing older history into learned summary embeddings that are not human-readable ^17^, in CLIN-SUMM the same durable longitudinal patient summary can be computed once and reused across endpoints, whether for manual clinical review, medication or vital sign extraction, symptom trend analysis, development of any number of machine learning models for disease classification or risk prediction, or other targeted tasks. This decouples representation construction from prediction, reducing repeated work during model development. Computational expense is also dramatically reduced, both because the summaries must only be computed once, and because the substantially compressed summaries require a fraction of tokens.

CLIN-SUMM uses a two-prompt architecture (one for baseline synthesis and another for incremental updates), automated redundancy filtering, and a sliding window for context management. The method is agnostic to the exact choice of LLM, and here we demonstrate CLIN-SUMM implemented with the open-source, low-cost Qwen3 ^18^ model, which ensures reproducibility and privacy-preserving local deployment. We evaluate the framework using longitudinal primary care data from a cohort of more than 12,000 patients and evaluate the summaries according to examples of their intended use, rather than relying solely on surface-level similarity to a reference text ^19,20^. We demonstrate that CLIN-SUMM summaries (1) achieve high compression efficiency, (2) maintain clinical fidelity under structured clinician review, with high scores for correctness and completeness, (3) underpin strong downstream performance in dementia diagnosis and 3-year prediction modeling, and (4) facilitate automated medication extraction. Together, these results suggest that incremental, structured longitudinal summarization can provide a scalable and reusable mechanism for unlocking the value of unstructured clinical notes for both patient care and predictive modeling.

## 2 Results

### 2.1 Summary generation and experimental setting

CLIN-SUMM longitudinally summarizes fragmented and repetitive EHR patient visit notes into a coherent, organized, efficient and date-partitioned summary representation for each patient that grows over time as clinical notes accumulate from encounters with the healthcare system. CLIN-SUMM uses a two-prompt architecture to convert clinical notes to compressed summaries using an LLM, as shown in **Figure 2**, with full prompt templates and technical implementation details provided in **Supplementary Figure 1** and **Supplementary Section 1 (Prompt Optimization and Parameter Selection)**. The first <u>initial summary prompt</u> uses an LLM to organize all of the text from the first clinical note into seven sections (“Chief Complaints”, “History of Present Illness”, “Past Medical History”, “Medications & Allergies”, “Vital signs or Lab/Imaging Findings”, “Diagnosis”, and “Treatment Plan and Follow-up”). The second, <u>change-focused update prompt</u> operates on each subsequent note. This prompt inputs the patient’s previously generated summaries so that the LLM adds only novel information from that specific encounter to each section and also populates a “Changes over Time” section that summarizes key temporal updates such as medication adjustments, new diagnoses, or changes in lab/imaging findings. CLIN-SUMM includes redundancy control via a Jaccard similarity ^21^ filter that avoids preprocessing near-identical notes, uses a sliding window mechanism to handle patients with long encounter histories without exceeding LLM context limits, and has a generalizable design that can be applied to a wide range of phenotypes and downstream tasks. Our setup is generic, and we demonstrate our main results using the Qwen3 ^18^ LLM.

**Figure 2:**
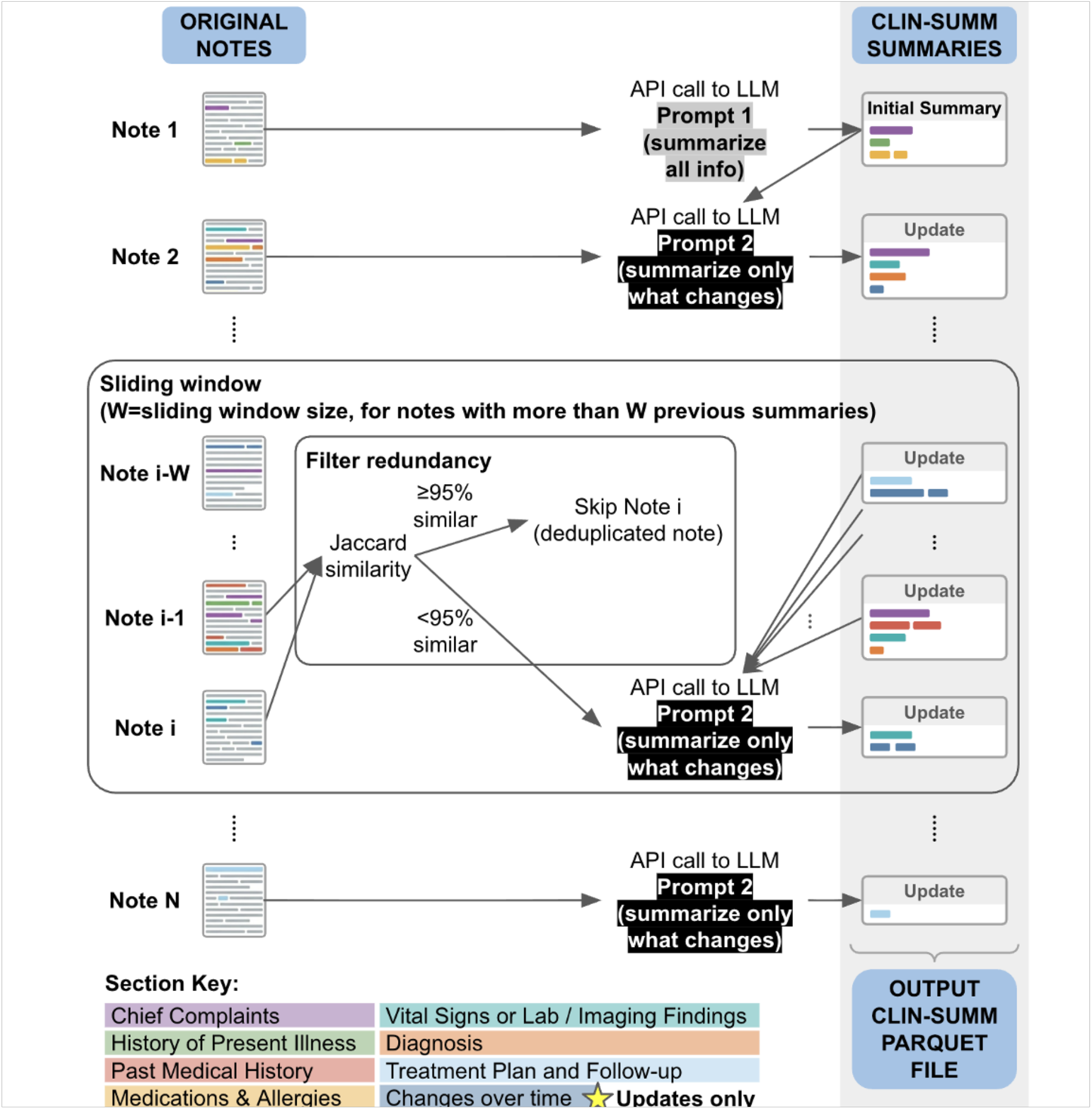
Raw longitudinal clinical notes are converted into structured patient timelines through sequential LLM-based summarization. The first note is summarized to create an initial patient summary, while subsequent notes are processed to extract only newly documented information. Redundant notes are filtered via a Jaccard similarity threshold, and a sliding-window mechanism limits context length for long note histories. The resulting updates are appended to a structured CLIN-SUMM representation, yielding a compressed, organized, date-partitioned longitudinal clinical summary for each patient.

We developed the methods, models and analyses using clinical notes from a matched case-control cohort drawn from the Primary Care Longitudinal Cohort (PCLC), which includes adult patients receiving longitudinal care at Massachusetts General Hospital (MGH) primary care practices between 2005-2023. Our cohort includes 6,178 dementia patients and 6,178 age- and sex-matched controls, with a total of 2,794,644 notes with an average of 15 years of follow-up per subject. Supplementary Table 1 describes the basic participant characteristics of the population. We used a pilot set of 1500 subjects (750 randomly selected dementia patients aged 50-75 at diagnosis and their corresponding 750 age- and sex-matched controls) for initial development. We subsequently used the entire cohort to evaluate the clinical utility of CLIN-SUMM summaries. Cohort selection, matching procedure, and data splits are described in detail in Methods Sections 4.1 and 4.5.1.

### 2.2 CLIN-SUMM enables significant compression

CLIN-SUMM summarization substantially compressed the clinical narratives in the full cohort, reducing 2,531,666,623 words (3,375,555,497 tokens) to 643,413,669 words (857,884,892 tokens), yielding an overall space savings of 74.59% (space savings is defined as the relative reduction in word count between the original clinical notes and their corresponding summaries). Analyzing compression at the patient level (which reduces the impact of patients with disproportionately numerous or lengthy notes) yielded a space savings of 57.8 ± 26.7%, indicating consistent compression across patients.

Early notes typically showed lower compression due to higher content density, while later notes were more highly compressed as CLIN-SUMM removed repeated information (**Supplementary Figure 2**). Compression was further evaluated at the patient level by analyzing the distribution of space-saving percentages. To account for multiple notes per day, notes were aggregated at the day level before computing average space savings per individual. While most patients showed positive compression, a small subset exhibited negative compression, more commonly in cases with very short notes (<200 words) or minimal new information. In these instances, the summarization process sometimes erroneously incorporated information from prior summaries, leading to longer outputs. These cases were often associated with low information note types (e.g., phone calls, emails, addenda), and 57% of notes in this group were under 200 words, suggesting limited information content as a key driver (**Supplementary Figure 3**).

### 2.3 CLIN-SUMM summaries achieve high correctness and completeness scores

A clinician-led review of CLIN-SUMM summaries from 698 notes (30 patients) sampled from the pilot set (*n* = 1,500, described above) using a structured questionnaire (**Supplementary Table 2**) to evaluate individual summaries at the note-level revealed high scores for correctness and completeness and low hallucination rates. An independent evaluation by a second clinician of summaries from 403 notes (15 patients) revealed high inter-rater agreement (**Supplementary Table 3**).

Correctness scores (assessing alignment of the summary with the key points in the original notes without factual errors or misinterpretations) averaged 4.69 ± 1.02 out of 5 with an inter-rater agreement of 88.8% [95% CI 86.8 - 90.4], with Cohen’s **κ** = 0.63. Completeness scores (assessing the level of information coverage in the summary compared to the original notes) averaged 4.65 ± 1.02 out of 5 with an inter-rater agreement of 87.1% [95% CI 85.4 - 88.6], with Cohen’s **κ** = 0.60. Dichotomizing scores into adequate (4-5) and inadequate (1-3) yielded Cohen’s **κ** values of 0.68 and 0.63, respectively **(Supplementary Table 4, Supplementary Figure 4)**.

Fewer than 4% of summaries exhibited hallucinations. Qualitatively, these mostly involved (1) inferring diagnoses that were not explicitly documented based on presenting symptoms, (2) misinterpreting vitals or laboratory values due to inconsistent formatting, and (3) incorrectly attributing lack of treatment to unavailability. Representative examples are provided in **Supplementary Section 2 (Error Analysis of CLIN-SUMM Summaries)**. These error types reflect known limitations of current LLMs and are likely to diminish as models continue to improve in reasoning, factual consistency, and clinical grounding.

To assess potential error accumulation across sequential summaries, we compared correctness, completeness, and hallucination rates across patient-specific early, middle, and late stages. No significant early-to-late differences were observed in correctness (p = 0.59), completeness (p = 0.96), or hallucination rates (p = 0.86). However, hallucinations were more likely in a subsequent summary when the preceding summary contained a hallucination (18.2% versus 2.3%; risk ratio = 7.83, 95% CI 2.43–17.32), suggesting possible temporal clustering of errors without progressive deterioration in overall summary quality (**Supplementary Figure 5; Supplementary Table 5**).

### 2.4 CLIN-SUMM summaries can form the basis for machine-learning modeling for early disease screening

#### 2.4.1 Dementia diagnosis and 3-year prediction

The rich textual data contained in patients’ historical clinical notes is summarized in the CLIN-SUMM longitudinal dataset and can be leveraged for machine learning. We demonstrated this by training transformer models for dementia diagnosis and 3-year prediction that input CLIN-SUMM summaries (and not the original, much longer clinical notes). The two models are based on Clinical ModernBERT ^22^, and are predictive, encoder-only models for binary classification of dementia cases versus controls. Note that the controls were not healthy: many had multiple comorbidities and overlapping clinical risk factors, making the task of distinguishing dementia cases versus controls more challenging.

The <u>diagnosis</u> model aims to improve dementia case identification by complementing International Classification of Diseases (ICD) and Systematized Nomenclature of Medicine (SNOMED)-based definitions, using notes to detect individuals who may not have formal codes. The <u>3-year prediction</u> model aims to identify patients at high risk of developing dementia, supporting opportunities for early, proactive intervention through preventative measures, lifestyle changes, and therapeutic strategies, potentially transforming how chronic diseases are managed. The diagnosis model was trained using all of each patient’s CLIN-SUMM summaries up to 30 days before formal dementia diagnosis, and the 3-year prediction model was trained using all of each patient’s CLIN-SUMM summaries up to 3 years before formal dementia diagnosis (**Figure 3A**). For controls, we used 30 days or 3 years before the censoring date, respectively. For the diagnosis model, the 30 day cutoff excludes periods likely to contain statements around the time of formal diagnosis, while still retaining less definitive references (e.g., concerns or evaluations for cognitive decline) or documentation of clinically suspected dementia that is not yet coded, thereby encouraging the model to rely on dementia-consistent clinical language patterns rather than direct statements of disease presence. In contrast, the 3-year prediction model aims to capture subtle early signals that might signal disease onset well before symptoms emerge. For very long summary histories, we truncated the input to the most recent part, corresponding to the period closest to the diagnosis date (or censoring date for controls), that could fit within Clinical ModernBERT’s context window (8,192 tokens). This strategy prioritizes information closest to the first recorded diagnosis or matched control reference date; it does not necessarily preserve the earliest documented history. At the three-year cutoff, 73.0% of case summaries and 68.7% of control summaries required truncation. Retention varied with note volume, and patients with dense records lost more early history (**Supplementary Tables 6–7; Supplementary Figure 6**). We also compared CLIN-SUMM summaries with raw clinical notes using the same Clinical ModernBERT architecture and 8,192-token limit, and also evaluated TF-IDF ^23^ models on the same selected text. The comparison methods and results are reported in **Supplementary Section 3 and Supplementary Table 8**.

**Figure 3:**
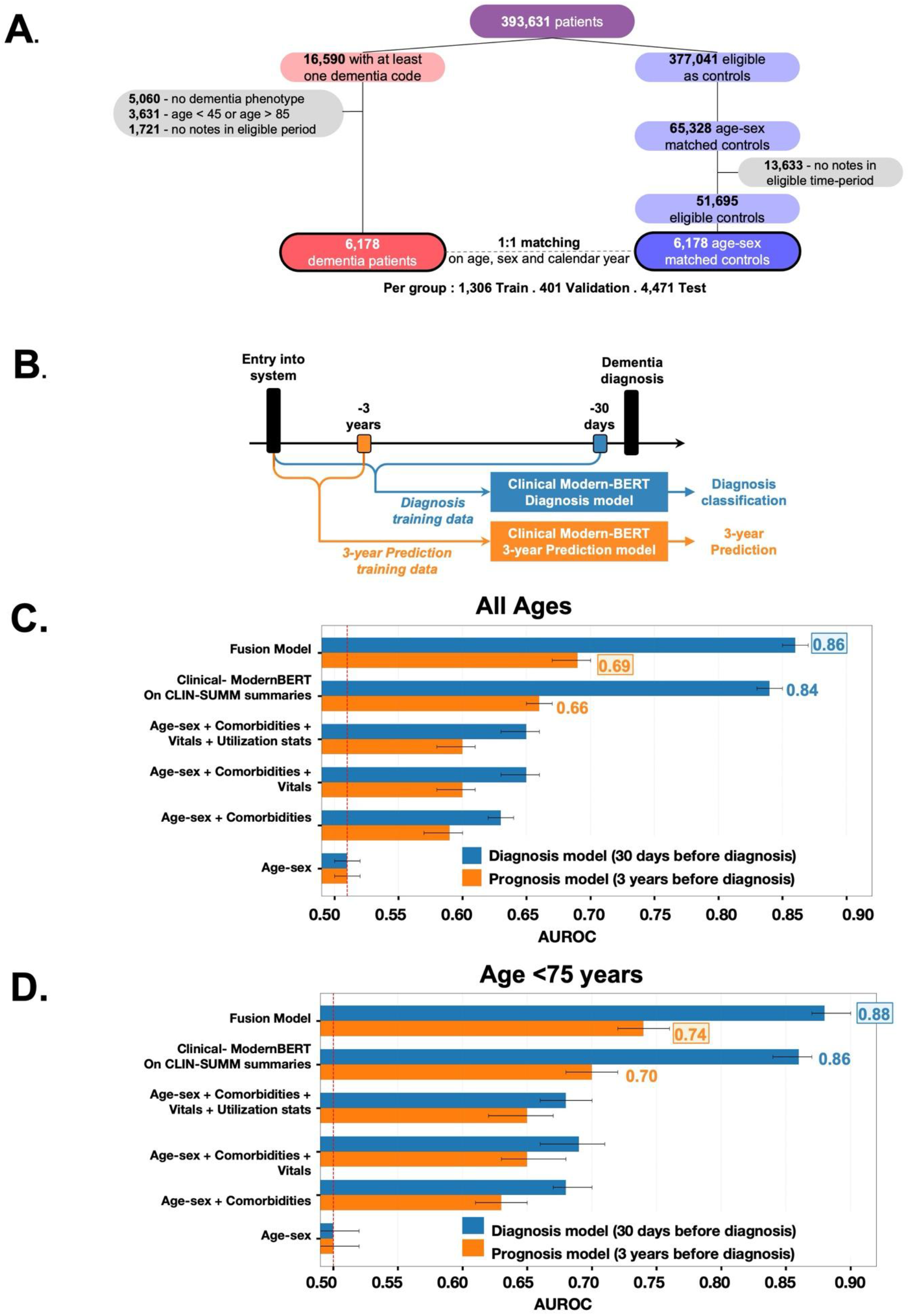
Overview of study design and modeling framework for dementia diagnosis and 3-year prediction. **(A)** Cohort construction and study flow diagram, controls were matched on age, sex, and calendar year. **(B)** Temporal framing of the prediction tasks, with diagnosis prediction evaluated 30 days before diagnosis and 3-year prediction evaluated three years prior to diagnosis. **(C-D)** Comparison of performance across models for all ages **(C)** and among people younger than 75 years **(D)**. The fusion model combines CLIN-SUMM prediction scores with age, sex, comorbidities, vital signs, and utilization measures and was evaluated in the subset with all required structured data; Clinical ModernBERT on CLIN-SUMM summaries model was evaluated on the full test set for each task.. Bars show AUROC, with the dashed red line indicating the age-sex baseline.

Clinical ModernBERT models using CLIN-SUMM summaries to identify dementia cases under the diagnosis and 3-year prediction scenarios consistently outperformed multifactorial dementia risk models ^24^ derived from the CAIDE dementia risk model ^25^ (**Figures 3B and 3C**, with detailed results in **Supplementary Table 9**). For the dementia diagnosis task, performance was strong (Area Under the Receiver Operating Characteristic (AUROC) 0.84, Area under the Precision-Recall Curve (AUPRC) 0.86, F1 score 0.75, precision 0.82, recall 0.69), reflecting high discrimination and a balanced trade-off between sensitivity and specificity. Because the held-out cohorts were approximately balanced between cases and controls, precision and AUPRC describe performance in this matched evaluation setting and should not be interpreted as expected performance at the lower dementia incidence of an unselected clinical population. As expected, the dementia 3-year prediction model had lower performance (given that the indicators of future dementia diagnosis are less specific the further one goes back in time and may be prone to misclassification) but retained sufficient discriminatory power (AUROC 0.66, AUPRC 0.67, F1 score 0.58, precision 0.64, recall 0.54). For context, logistic baselines incorporating age, sex, prevalent comorbidities and vitals (CAIDE dementia risk model), showed more limited predictive ability in the subset of test subjects with vitals (diagnosis: AUROC 0.65 vs. 0.86; 3-year prediction: AUROC 0.60 vs. 0.68), reflecting the insufficiency of structured risk factors alone in this setting. These limitations may in part be attributable to incomplete capture of clinical nuance (e.g., symptom progression, cognitive complaints, and clinician assessments), variability in data availability (e.g., missing or irregularly recorded vitals), and constraints in representing longitudinal context using structured features (e.g., temporal ordering and evolution of clinical events). We also evaluated a 3-year prediction fusion model combining the CLIN-SUMM summary prediction scores with age, sex, comorbidities, vital signs and utilization measures. Among 4,761 patients with the required inputs, the fusion model achieved an AUROC of 0.69 (95% CI, 0.67–0.70) and an AUPRC of 0.68 (0.66–0.70), with detailed results in **Supplementary Table 9**. Discrimination was stronger among patients younger than 75 years (*n* = 1,753; AUROC 0.74, 95% CI 0.72–0.76; AUPRC 0.74, 0.71–0.77) than among those aged 75 years or older (*n* = 3,008; AUROC 0.66, 0.64–0.67; AUPRC 0.65, 0.62–0.68) (**Figure 3D**).

Identifying elevated risk earlier in life may provide more time for clinical assessment, follow-up and potential intervention. Cases had more eligible notes and available summary tokens than controls at both cutoffs (**Supplementary Table 10**). We therefore evaluated how well documentation and utilization variables could distinguish cases from controls. For diagnosis, a model using eligible note count, available history duration, and summary token count achieved an AUROC of 0.55 (95% CI, 0.54-0.56), compared with 0.84 (0.83–0.85) for Clinical ModernBERT on summaries. For 3-year prediction, the corresponding AUROCs were 0.54 (0.52-0.55) and 0.66 (0.65-0.67) (**Supplementary Table 11**). These measured utilization variables did not reproduce the summary model’s discrimination, although other aspects of healthcare use may contribute to its predictions.

The endpoint for both tasks was the first recorded dementia diagnosis, rather than disease onset. The 30-day diagnosis model assessed discrimination shortly before that date, when symptoms, cognitive assessments, medication use, or diagnostic language might already be documented. To assess sensitivity to proximity to diagnosis, we retrained the models using 90-day and six-month cutoffs instead of 30 days. AUROC was 0.84 (95% CI, 0.83-0.85) at 30 days, 0.82 (0.82-0.83) at 90 days, and 0.78 (0.77-0.79) at six months (**Supplementary Table 12**). These results indicate stronger discrimination nearer the recorded diagnosis, while showing predictive signal at earlier cutoffs.

#### 2.4.2 Early detection time horizon

To evaluate the model’s clinical utility and early-detection capabilities, we conducted a longitudinal lead-time analysis. We applied the frozen diagnosis model (originally trained using all CLIN-SUMM summaries up to 30 days before diagnosis) to incrementally expanded temporal windows, simulating deployment in a real clinical scenario in which a risk assessment model is repeatedly applied as a patient’s clinical trajectory evolves (**Figure 4A**). Detailed longitudinal evaluation procedures are provided in **Supplementary Section 4 (Early detection time horizon).**

**Figure 4:**
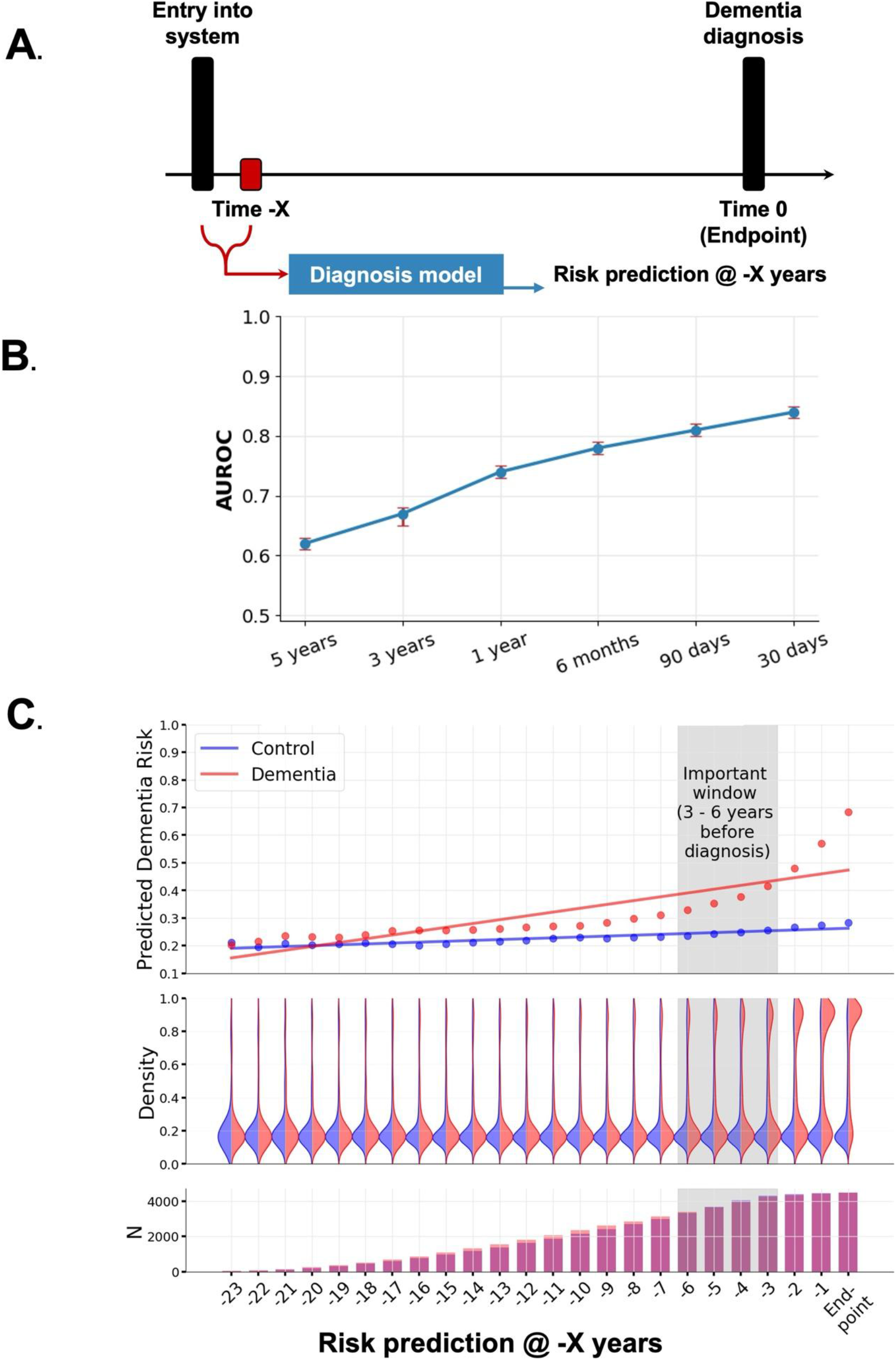
Longitudinal dementia risk prediction across time prior to diagnosis. **(A)** Temporal prediction framework illustrates how the diagnosis model estimates dementia risk at different time horizons before the diagnostic endpoint (Time 0). **(B)** Diagnosis model performance across prediction windows, showing AUROC for risk prediction evaluated at 5 years, 3 years, 1 year, 6 months, and 30 days prior to diagnosis, with performance increasing as predictions approach the diagnosis date. **(C)** Predicted dementia risk across time windows for dementia cases and controls. Mean predicted risk diverges between dementia cases and controls as the endpoint approaches, with a pronounced divergence **3–6** years before diagnosis. The middle panel shows the distribution of predicted risks for both groups across time, and the bottom panel shows the number of samples available at each prediction window.

When applied to test sets right-truncated at progressively earlier time points (−5 years, −3 years, −1 year, −6 months before diagnosis), the diagnosis model generalizes to an early prediction tool (**Figure 4B**, with detailed results in **Supplementary Table 13)**. The model maintained meaningful discriminatory power even years before diagnosis, with an AUROC of 0.78 at −6 months and 0.74 at −1 year. Notably, the 30-day diagnosis model achieved an AUROC of 0.67 on data truncated at −3 years, closely matching the 3-year prediction model (AUROC 0.66) despite significant differences in the training data.

Longitudinal patterns in mean predicted dementia risk, computed over progressively longer CLIN-SUMM summaries in one-year increments, revealed differences in the dementia vs control groups and suggest that model predictions are grounded in clinically meaningful cues that evolve over time (**Figure 4C**). Many years prior to dementia diagnosis (or for controls, the censor date), the dementia and control groups had similar mean predicted dementia risk, both of which were below 0.2. The mean dementia risk steadily increased in the dementia group as the clinical histories became progressively richer and the formal dementia diagnosis dates neared, while the mean dementia risk in the control group was stable. A visible separation between the distributions of predicted dementia risk for the dementia versus controls groups is clear between 3 to 6 years before diagnosis, consistent with our previous findings of high AUROC for binary classification. Plotting mean dementia risk on a two-dimensional grid as a function of both the number of years prior to endpoint and subject age at the time (**Supplementary Figure 7**) reveals that dementia risk generally increases with age so long as the endpoint is sufficiently near (e.g., a maximum of 6 years away), but that differences in mean predicted dementia risk between the two groups are highest near clinical endpoints regardless of age.

#### 2.4.3 Interpretability

To understand which linguistic features drive model predictions, we analyzed word-level attribution scores derived from LIME ^26^ explanations of the fine-tuned Clinical ModernBERT dementia diagnosis model (**Figure 5**). Across semantic categories, words associated with cognitive impairment and neurological symptoms were among the words with the highest positive attribution scores, indicating strong positive contributions to dementia predictions. These included terms such as ‘cognitive’, ‘memory’, ‘impairment’, ‘confusion’, ‘decline’, ‘deficits’ and ‘neuropsychological’ ^27,28^. Additional promoting signals included functional or symptom descriptors such as ‘fatigue’, ‘fall’, ‘distress’ and ‘instability’, which are commonly documented in patients with neurocognitive disorders. In a sensitivity analysis using Integrated Gradients ^29^ on Clinical ModernBERT’s native subword tokens, 77.8% of the prespecified Figure 5 features had the same attribution direction as in LIME, with a Spearman rank correlation of 0.74 (**Supplementary Figure 8**). The 34 features with discordant directions generally had attribution scores near zero under both methods. Thus, the broader patterns were reasonably consistent across methods, while attribution of weaker features was less stable. Given that LIME operates on whitespace-tokenized words, certain multi-word clinical concepts (e.g., ‘hearing loss’) may appear as separate tokens (e.g., ‘hearing’ and ‘loss’, both of which were identified as promoting words). In exploratory analyses, co-occurrence patterns identified via simple regex-based matching showed that ‘hearing’ and ‘loss’ appeared together within the same context in 9 of 17 individuals (52.9%) among those in whom either term was identified. This co-occurrence is consistent with references to hearing loss, a known risk factor for cognitive decline and dementia ^30^, but the separate LIME attributions do not establish that the model attributed its prediction to the combined clinical concept. Additional words associated with established risk factors and comorbidities, including depression and hypertension, also showed positive attribution, consistent with their links to increased dementia risk ^30–32^. In contrast, words with negative attribution scores, contributing toward the control class, were largely associated with routine clinical documentation, stable clinical status and general health descriptions. These included terms such as ‘maintenance’, ‘screening’, ‘care’, ‘annual’, ‘improving’ and ‘managing’. Overall, the salience patterns demonstrate that the model prioritizes clinically interpretable signals consistent with known manifestations of dementia, including cognitive impairment, neurological symptoms and functional decline. These findings suggest that the model’s predictions are driven by medically relevant language rather than spurious textual artifacts. Because the analysis sampled only the extremes of predicted risk, these patterns do not characterize explanations across the full risk distribution nor establish that the model relies on these terms for every patient.

**Figure 5:**
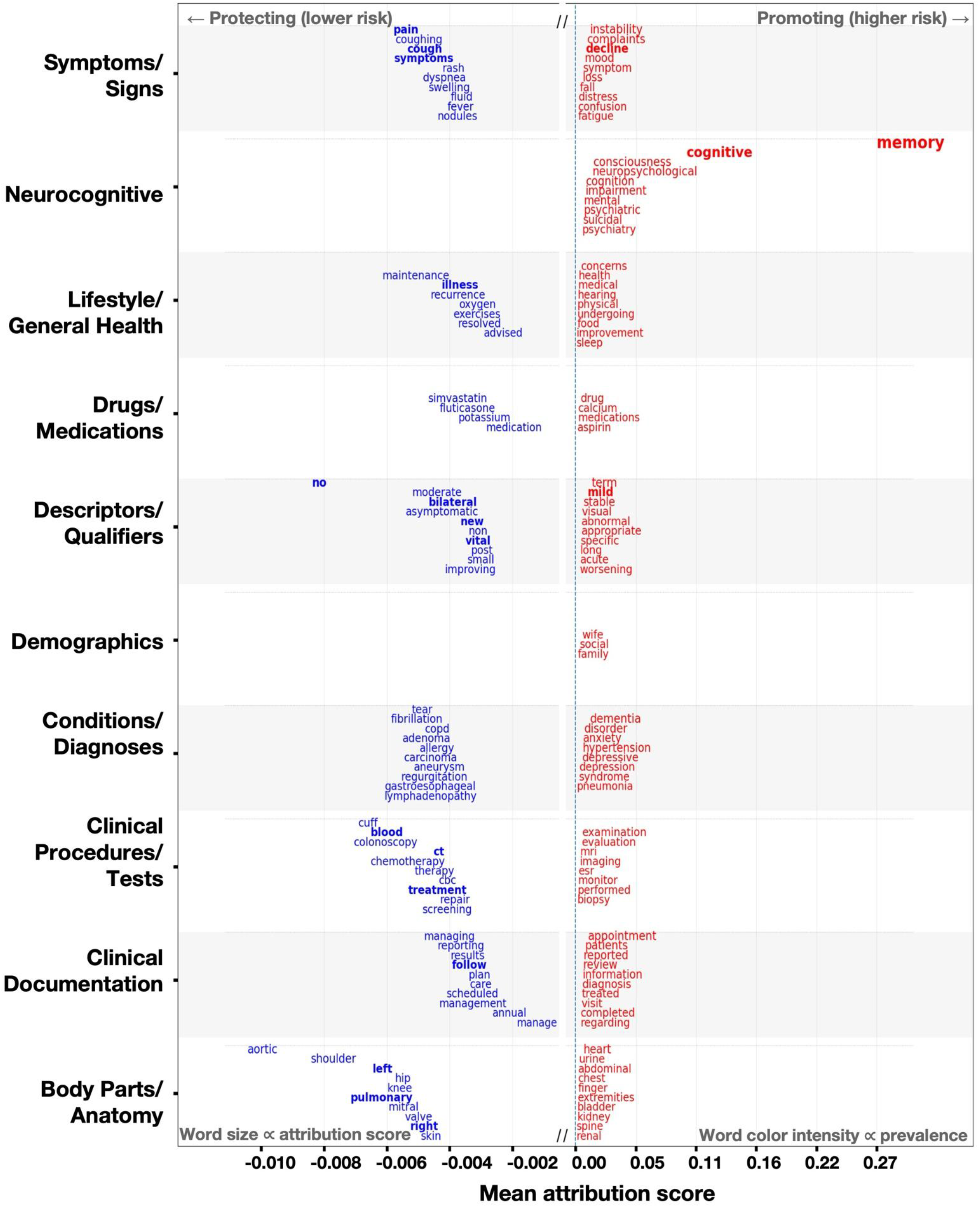
Word-level attribution scores derived from **LIME** highlight influential features contributing to dementia risk in the diagnosis model. Word size and position reflect attribution magnitude, and color intensity reflects prevalence in the dataset. Blue terms correspond to features associated with lower predicted risk, while red terms indicate features associated with higher predicted risk. **Neurocognitive** terms and **clinical diagnoses** show strong positive attribution, whereas general health and procedural terms tend to show negative attribution.

#### 2.4.4 Additional analyses of model behavior

Additional analyses examining model behavior, including error patterns and evaluation of predictions in relation to mild cognitive impairment (MCI), are provided in **Supplementary Section 5 (Additional analyses of model behavior) and Supplementary Figure 9**. Overall, misclassifications were primarily associated with ambiguous or incomplete clinical context, while analyses of MCI-related subgroups showed graded risk patterns consistent with disease progression.

### 2.5 CLIN-SUMM summaries augment medication extraction

As medication information is consolidated into a dedicated section for each patient summary, efficient extraction is possible by querying a single structured segment of the longitudinal record, eliminating the need to scan thousands of raw clinical notes. To assess the completeness of medication extraction, we compared Donepezil medication events extracted by GPT-4o through the OpenAI API from locally generated Qwen3 CLIN-SUMM summaries, with medication records available in structured EHR tables (Snowflake). Because structured records were available only from 2015 onward and could extend beyond the last note included in a summary, we restricted the source comparison to each patient’s shared observation window: from 2015 through the earlier of the last included note or censoring date (**Figure 6A**). This comparison served both as a validation step and as a test of the hypothesis that summary-based extraction may reduce apparent missingness in structured data sources. Within the shared observation window, CLIN-SUMM identified 1,550 Donepezil events (1,520 in dementia patients; 30 in controls), and structured EHR data identified 1,275 (1,235 in dementia patients; 40 in controls). Across both groups, 1,211 events were found in both sources, 339 only in CLIN-SUMM and 64 only in structured EHR data. We reviewed 100 of the 339 CLIN-SUMM-only events in the shared observation window against their source notes; 94% were supported as documented medication use, 5% reflected medication information in the summary that was unsupported by the source notes, and 1% resulted from errors when the extraction LLM queried the summaries. We also manually reviewed 40 of the 64 structured-only events. For 23 (57.5%), no note was available on the structured event date, and surrounding notes contained no mention of the medication event. Fourteen events involved notes documenting medication discontinuation, for which GPT extracted a dose of 0; 12 of these notes postdated the structured medication event, and two occurred on the same day. In one event, the summary mentioned Aricept but omitted its dose, which had been extracted from an earlier summary slice before 2015; the source note indicated no change in treatment. In another event, the summary mentioned the medication without a dose. In the remaining event, the summary omitted the medication entirely. Source-note verification of CLIN-SUMM-only events supports the value of summary-based extraction for recovering medication information documented in notes but absent from the available structured data. Gaps in note coverage may explain some structured-only events, although event counts alone do not establish the completeness of either source. To further evaluate concordance between sources, we matched the doses of CLIN-SUMM extracted medication events with the structured medication record per patient within the shared observation window, treating brand and generic names as equivalent. We first chose the pair with the smallest difference between the summary event date and structured medication order date. For 19 of 1,211 events (1.6%), we selected an alternative pair with matching doses whose date gap was no more than two days greater than the minimum, accounting for multiple strengths recorded on the same or nearby dates. Dosage agreement between CLIN-SUMM extractions and structured records was high, with 96.0% of events showing exact agreement. Discrepancies were infrequent and largely explained by clinically interpretable factors, including dose titration (1.9%), where notes captured gradual dose escalation while structured records reflected the final prescribed dose, and split or fractional dosing (0.3%), where notes documented detailed administration patterns (e.g., ¼ tablet taken twice daily), while structured records capture only the prescribed medication without encoding these instructions. In 0.5% of cases, the CLIN-SUMM extracted dose aligned with the structured ‘description field’, while the structured ‘dose quantity’ field was inaccurate, highlighting inconsistencies within structured medication data. True mismatches were observed in 1.3% of cases, although the raw clinical notes supported the CLIN-SUMM extracted dose, indicating inconsistencies in structured records. The discrepancy categories were mutually exclusive, and all percentages were calculated using the total number of selected matched pairs as the denominator. Together, these results suggest that CLIN-SUMM summaries may improve medication capture while maintaining strong agreement with the clinically administered dosage.

**Figure 6:**
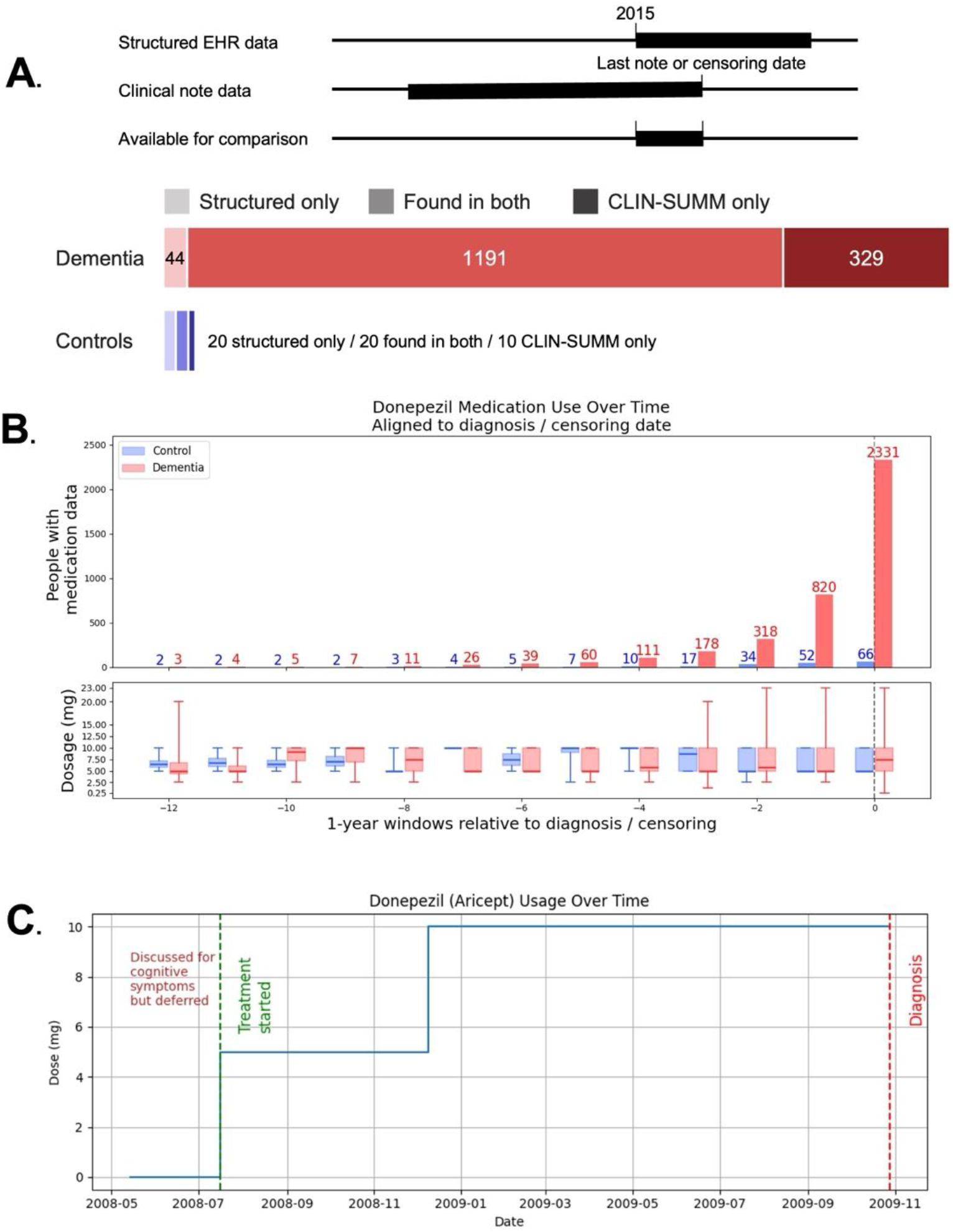
Medication extraction and longitudinal treatment patterns. **(A)** Comparison of medication extraction counts from CLIN-SUMM summaries and structured EHR data within each patient’s shared observation window, from 2015 through the earlier of the last included note or censoring date. **(B)** Population-level trends in **Donepezil** use and dosage in dementia patients and controls, derived from all available CLIN-SUMM-extracted events and aligned relative to dementia diagnosis or control censoring date. **(C)** Example patient trajectory illustrating medication initiation and dose changes over time.

Medication trajectories reconstructed from CLIN-SUMM summaries revealed clinically meaningful treatment patterns for dementia-related medications. For example, Donepezil use increased substantially in the years preceding diagnosis among dementia patients, with the number of individuals receiving the medication rising sharply near their diagnosis dates (**Figure 6B**). In contrast, Donepezil use remained rare among controls across the observation period. This likely reflects prescriptions for suspected cognitive impairment or early memory complaints that did not lead to a recorded dementia diagnosis during the study period, or individuals who may have received a diagnosis after the study cutoff date, or lagging documentation practices following diagnosis of dementia. Dosage distributions (**Figure 6B**) also showed increasing variability closer to diagnosis in the dementia cohort, consistent with treatment initiation and adjustment during disease progression, while trajectories for individual patients show trends over time (**Figure 6C**).

Together, these results demonstrate that CLIN-SUMM summaries can serve as an important source for reconstructing longitudinal medication trajectories and may help mitigate missingness, inconsistencies, fragmentation or lag in structured EHR medication data with reduced computational cost compared to processing the original notes.

### 2.6 Evaluation of a Commercial Frontier LLM

Although CLIN-SUMM is designed to operate with any LLM, and Qwen3 ^18^ was ultimately used for summarization in this study, we explored a state-of-the-art proprietary LLM as a reference point given that such models are widely regarded as benchmarks for high-quality text generation. Specifically, we generated CLIN-SUMM summaries using the GPT-4o ^33^ LLM in the pilot set. We then evaluated how similar Qwen3-generated summaries were to GPT-40-generated summaries to support cost-efficient and scalable deployment across the full cohort. Qwen3 produced substantially similar summaries, with a mean BERTScore recall of 0.91 between Qwen- and GPT-generated text (**Supplementary Table 14)**. BERTScore compares generated text; it does not measure factual accuracy against the source notes or establish equivalent downstream predictive performance. These findings demonstrate that open-source alternatives such as Qwen3 can generate similar longitudinal summaries of clinical notes as proprietary LLMs, while substantially reducing computational and financial costs (e.g., $250 with GPT vs. $35 with Qwen3 for summarizing 100 individuals from our cohort with an average of 100 notes each), and mitigating key barriers to deployment, including limited scalability and restricted accessibility associated with application programming interface (API)-based models. The reported costs are estimates based on API prices and compute rates at our institution at the time of analysis, as well as the number of input and output tokens processed in this dataset. Costs will vary with note volume, text length, generated summary length, and changes in pricing. Moreover, since Qwen3 can be deployed entirely within a secure, institution-controlled environment, it eliminates data-transfer risks and enhances compliance with privacy regulations, including HIPAA (Health Insurance Portability and Accountability Act), an essential requirement for clinical translation.

### 2.7 Additional Applications

Additional pilot applications illustrating the versatility of CLIN-SUMM summaries, including applications in disease progression analysis and longitudinal symptom trend characterization, are presented in **Supplementary Section 6 (Additional Applications of CLIN-SUMM Summaries)**.

## 3 Discussion

In this study, we introduce CLIN-SUMM, a longitudinal summarization framework designed to condense clinical notes and create a reusable representation of narrative EHR data that preserves temporal evolution while reducing redundancy. Rather than repeatedly processing raw, unstructured notes for each downstream task, CLIN-SUMM produces a structured, date-partitioned representation that captures incremental clinical updates over time. This representation retains the nuance, versatility, and comprehensibility of raw clinical text while overcoming its inherent redundancy and lack of organization, and can be incrementally updated as new encounters occur, avoiding the need to regenerate summaries from scratch in clinical workflows. In doing so, CLIN-SUMM defines a new intermediate layer between raw clinical text and downstream applications, offering a more tractable and scalable way to reason over longitudinal patient trajectories. Compared to existing approaches, this representation offers distinct advantages over structured codes ^1–3^ (which often suffer from missingness and coding errors, and lack nuance), original notes ^15,16^ (which are lengthy, repetitive and unorganized), hierarchical textual representations ^17^ (which are derived from notes and structured data and retain many of their problems), learned summary embeddings ^17^(which are not human-readable for clinical tasks) and conventional clinical summaries ^10–13^ (which typically lack an explicit temporal dimension, although recent research has highlighted the benefits of summarizing on a daily basis ^34^).

From a clinical perspective, this representation enables a more efficient and interpretable view of patient history and how it may evolve over time. By isolating only the “new” clinical signal at each encounter, CLIN-SUMM reduces the cognitive burden associated with manual chart review and allows clinicians to rapidly grasp longitudinal changes. The structured, temporally organized summaries support tracking of evolving problems and medications, can be updated as new notes are processed, and provide a compact view of documented clinical changes that may help clinicians review activity since a patient’s previous visit. High clinician-rated scores for correctness and completeness, alongside a low hallucination rate, suggest that these summaries can serve as a reliable cognitive scaffold in clinical workflows, for example by providing an update of relevant clinical activity since a patient was last seen by a clinician. Importantly, CLIN-SUMM can be generated using open-source models in a local, privacy-preserving setting, allowing institutions to create the longitudinal representation within their own environment. The medication extraction analysis further highlights a separate downstream use of that representation: while some medication events overlap with structured EHR tables, many are captured only within note-derived summaries, indicating that CLIN-SUMM complements and augments structured data sources. For this analysis, we used GPT-4o through the OpenAI API; the summaries’ reduced length limited the text sent to the API and its associated token costs.

Beyond clinical applications, CLIN-SUMM introduces a new paradigm for machine learning on clinical text by decoupling raw note processing from prediction. As an application-agnostic representation, CLIN-SUMM summaries can be generated once at the system level and reused across multiple downstream tasks, eliminating the need to repeatedly process raw notes and improving computational efficiency. For example, the same set of summaries could support models for predicting or diagnosing a range of conditions, such as diabetes, cardiovascular disease, stroke, and dementia, without requiring task-specific reprocessing of the underlying clinical notes. The substantial reduction in token count further enables more scalable training and inference while preserving clinically meaningful signals. Given the heterogeneity of raw clinical text and variability in documentation practices, this abstraction layer represents a key step toward robust and generalizable machine learning systems. In this sense, the primary contribution of CLIN-SUMM extends beyond summarization to establishing a reusable infrastructure for longitudinal clinical reasoning and learning. Compression with clinical fidelity is not merely a technical metric, but a bridge to deployment: reducing information burden while preserving clinical accuracy makes CLIN-SUMM both usable and trustworthy in real-world settings. Moreover, by leveraging compressed representations instead of raw clinical notes, CLIN-SUMM reduces computational and financial costs, an important consideration for deployment at scale across large healthcare systems.

We evaluated this framework using diagnosis and future risk prediction of dementia as a proof-of-value application, motivated by the inherent difficulty of these predictive tasks due to the long prodromal phase of this disease and reliance on subtle, distributed signals in clinical notes. In our matched cohort, models based solely on demographic features (age and sex) performed near chance, indicating that signal from demographic factors alone is limited. Despite achieving substantial compression, CLIN-SUMM summaries supported robust classification performance for diagnosis and enabled meaningful future risk prediction up to three years in advance. This predictive performance compares favorably to established multifactorial models like CAIDE ^25^ and ANU-ADRI ^35^, yet it operates directly on existing notes without requiring structured variables that are often missing. These findings demonstrate that compression is not destructive: CLIN-SUMM preserves clinically relevant signals while removing redundancy, enabling effective prediction without reliance on raw notes or structured-only approaches.

Despite being trained on notes within 30 days of diagnosis, our dementia diagnosis model accurately distinguished dementia patients from controls up to 3–6 years in advance. Specifically, the model assigned approximately the same predicted dementia risk to dementia cases and controls many years prior to diagnosis (as early records likely contain fewer explicit disease-related signals and less frequent or subtler cognitive, neurological or behavioral indicators), and a steady separation between the two groups over time (as later notes capture evolving symptoms and correlated diagnoses). This capability is particularly vital given that dementia is insidious in onset, and formal ICD codes are often either never recorded or lag significantly. Indeed, in our cohort dementia medications were sometimes prescribed up to two years before a formal code appeared, yet the model accurately distinguished dementia patients from controls well before this lead-time. Importantly, intermediate-risk patterns observed in patients with MCI further support that the model is capturing a graded, longitudinal disease trajectory rather than simply detecting the presence or absence of explicit diagnostic factors.

Interpretability analyses suggest that the model is learning a clinically coherent signature of dementia rather than relying solely on heuristic cues or explicit diagnostic language. Notably, CLIN-SUMM appears to preserve clinically relevant information in a way that allows these signals to be effectively leveraged by the model. Beyond expected neurocognitive terms such as *memory* and *cognitive*, the model assigns importance to hearing-related language, dizziness, fall-related terms, mood changes, depression, and vascular or metabolic comorbidities, aligning with prior literature ^30–32^. These signals are best interpreted as evidence of clinical face validity rather than causal relationships, and they support the conclusion that CLIN-SUMM captures a distributed and realistic prodromal signature embedded in routine clinical documentation.

Error analysis further strengthens confidence in the model by revealing systematic and clinically interpretable failure modes. False positives are often observed in patients whose records contain dense cognitive-related vocabulary, while false negatives are more common in cases with sparse documentation or fragmented care across specialties. Importantly, discrepancies between model predictions and ICD labels often suggest potential label noise and delayed coding. In many such cases, model predictions appeared clinically plausible, supporting the reliability of the model in capturing meaningful clinical patterns.

While dementia serves as a compelling use case, the utility of CLIN-SUMM extends well beyond a single disease prediction task. The framework facilitates reconstruction of medication trajectories, tracking of symptom evolution, and summarization of disease progression over time. These capabilities support a broad range of applications, including longitudinal phenotyping and clinical decision support. We anticipate that CLIN-SUMM will be particularly valuable for conditions characterized by gradual and complex trajectories, such as autoimmune diseases, chronic kidney disease, and oncology treatment response, where relevant signals accumulate over time. This positions CLIN-SUMM as a general-purpose infrastructure rather than a task-specific solution.

Several limitations warrant consideration. First, although CLIN-SUMM demonstrated low hallucination rates, even rare factual errors carry clinical risk, and the sequential summarization framework may propagate or amplify errors over time. While prompt-level safeguards aim to constrain outputs to verifiable information, future work will focus on strengthening reliability through systematic error analysis, incorporation of source attribution to link summaries to original notes, and additional validation and monitoring strategies to ensure safe clinical integration. Technical refinements, such as optimized redundancy filtering, improved windowing strategies, and the pre-pruning of uninformative notes, could further enhance both summary accuracy and computational efficiency. Also, it is worth noting that the accuracy of the generated summaries and the cost required to compute them will improve alongside new developments in LLM technology. Second, even though training was restricted to notes prior to 30 days before diagnosis, potential prediagnostic signal leakage cannot be fully excluded. However, our longitudinal analyses and findings related to MCI suggest that model predictions are not solely driven by explicit diagnostic terms, supporting the presence of meaningful prediagnostic patterns. Third, documentation volume and healthcare utilization may contain useful predictive information and may also influence the language captured in summaries. The measured utilization variables alone had substantially lower discrimination than the summary model, but this comparison cannot exclude contributions from other aspects of healthcare use. Moreover, precision and AUPRC were measured in approximately balanced case-control test sets and may differ substantially in populations with a lower outcome prevalence. Fourth, direct comparisons with raw clinical notes under the same 8,192-token limit showed comparable or modestly better discrimination with CLIN-SUMM summaries. The summaries retained a longer span of patient history within that limit, so these comparisons reflect the combined effects of compression and longitudinal organization. We evaluated this approach in dementia; the value of retaining more history may differ across chronic, progressive, episodic, and acute conditions. Fifth, clinician evaluation was conducted on a limited sample due to the time-intensive nature of manual review, which requires clinicians to read, interpret, and reconcile information across numerous lengthy, technically complex clinical notes per patient and to verify alignment with the generated summaries. Expanded validation, either through larger-scale clinician assessment or complementary evaluation using LLM-based approaches, is needed for more robust and comprehensive assessment. Finally, external validation across diverse healthcare systems is necessary to assess generalizability and robustness across diverse patient populations, clinical settings, and documentation practices. In addition, the clinical context in which models trained on CLIN-SUMM summaries are applied may influence performance; for example, applying these models to patients seen primarily in specialty care rather than longitudinal primary care may increase the risk of false negatives due to more limited or focused documentation of certain clinical conditions.

Despite these limitations, CLIN-SUMM provides a practical pathway for transforming longitudinal clinical notes into a reusable infrastructure layer for both clinical reasoning and machine learning. By substantially reducing narrative volume while preserving temporal context and clinically meaningful signal, CLIN-SUMM enables more efficient chart review and supports robust downstream prediction, including early disease risk stratification years prior to diagnosis. As an application-agnostic representation, it can be computed once and reused across tasks, offering a scalable approach to leveraging unstructured clinical data. These findings position CLIN-SUMM as a promising foundation for integrating longitudinal clinical narratives into real-world analytical and decision-support systems.

## 4 Methods

### 4.1 Study Cohort and Data

We developed the methods, models, and analyses in this study using EHR data from individuals receiving longitudinal care at MGB primary care practices. The study population was derived from the **Primary Care Longitudinal Cohort** (PCLC), which includes adult patients linked to the Massachusetts General Hospital (MGH) primary care network between 2005 and 2023; their available longitudinal clinical records could extend earlier. The PCLC retrospectively identifies patients seen in primary care within the prior three years and assigns them to a primary care provider (PCP) or practice based on rules incorporating patient age, time since last visit, and visit patterns. Although longitudinal clinical data were available as early as 1976, patient–PCP linkage was assessed beginning in 2005.

PCLC comprises 393,631 patients and includes both structured and unstructured data. This includes patient demographics, diagnosis codes, procedure codes, medications, notes from health care encounters and annual ambulatory visit notes. Prior to downstream processing, all clinical notes were processed for Protected Health Information (PHI) minimization, including the removal or masking of identifiable information such as patient and provider names. As the machine learning models developed in this study focus on dementia diagnosis and 3-year prediction, we conducted a retrospective case-control study (**Figure 3A**). Subjects diagnosed with dementia were identified based on the presence of at least two relevant ICD or Systematized Nomenclature of Medicine (SNOMED) codes, which must have been recorded on different dates within a 12-month period (**Supplementary Tables 15 and 16**). In order to be included, subjects must have been aged 45-85 at their initial dementia diagnosis. To ensure a 3-year lead-time, we only included subjects who had notes documented prior to the 3-years window preceding the first dementia diagnosis code, yielding a final set of 6,178 dementia subjects. From this group, we randomly selected a pilot cohort of 750 subjects aged 50-75 years at their initial dementia diagnosis. An age-and sex-matched control group was selected from patients without any dementia-related codes, such that the sampling maintained a balanced 1:1 case-to-control distribution for the entire cohort. For each candidate control, we set the reference date to when they reached the matched case’s age at first recorded dementia diagnosis. We required this date to be within one year of the case’s diagnosis date, thereby aligning age and bringing birth year and calendar time into close correspondence. For 210 cases, the calendar-date tolerance was relaxed to three years to identify an eligible control. Matching was performed without replacement. The 30-day and three-year cutoffs were then applied to the respective case diagnosis and control reference dates. In total, the dataset included 1,605,524 ambulatory and visit notes from the dementia group (1,034,564 recorded prior to the dementia diagnosis date) and 1,189,120 ambulatory and visit notes from the control group (890,064 recorded prior to the censor date). The cohort spanned 870 primary care providers (PCPs) across 12,356 patients and included diverse note types and documentation templates (>100,000 templates, capturing variation across PCPs, nurses, and other care settings), reflecting substantial heterogeneity in clinical practice and documentation, even within a single-institution setting. Detailed demographic characteristics are described in **Supplementary Table 1**.

### 4.2 CLIN-SUMM Summarization

#### 4.2.1 Summarization Workflow

CLIN-SUMM uses two distinct prompts to iteratively summarize longitudinal clinical notes (**Supplementary Figure 1**). The initial prompt generates a comprehensive summary of the patient’s inaugural entry note into the system, with explicit sections encompassing “Chief Complaints”, “History of Present Illness”, “Past Medical History”, “Medications & Allergies”, “Vital signs or Lab/Imaging Findings”, “Diagnosis”, and “Treatment Plan and Follow-up”. The second prompt explicitly incorporates the previously generated summaries and aims to expand the patient-level summary with new information from each subsequent note, focusing exclusively on summarizing only novel content not encompassed within all of the preceding note-level summaries, and appending a “Changes Over Time” section that summarizes key temporal updates across encounters, such as medication adjustments, new diagnoses, or changes in lab/imaging findings, to track the patient’s evolving clinical state. Each summarized entry is timestamped using the original note’s date and time, preserving the chronological order of events.

CLIN-SUMM output is stored as a structured Parquet file, linking each patient’s notes with the note date and time, brief note-level descriptions (such as ‘patient note’, ‘wellness annual’, ‘lab results’, ‘medication-management’, ‘consult’ or ‘neurology’, provided by the EHR), and the generated summaries. This format preserves longitudinal detail while enabling flexible aggregation of summaries by month, year, clinical event, or section.

#### 4.2.2 Prompt Optimization and Parameter Selection

Prompts were iteratively refined following best practices ^36^, to produce structured, clinically faithful summaries. The CLIN-SUMM framework was implemented using the Qwen3 model ^18^ deployed via the vLLM inference framework ^37^. The approach incorporates context management and redundancy control mechanisms (via Jaccard similarity evaluation ^21^) to support stable longitudinal summarization. Full prompt templates, parameter settings and implementation details are provided in the Supplementary Section 1 (Prompt Optimization and Parameter Selection).

### 4.3 Compression Efficiency

We measured compression efficiency in 2,794,644 notes from the 12,356 patients in the full cohort. For each note, we compared the length of the original clinical text with the length of the corresponding CLIN-SUMM summary. Space savings is computed as the percent reduction in the number of words via

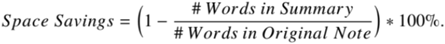

Compression was evaluated at multiple aggregation levels. First, overall space savings was computed across the entire cohort by comparing the total number of words in all original notes versus the generated summaries. Second, patient-level compression was computed by calculating space savings for each patient individually and summarizing the distribution across patients. Because multiple clinical notes may be generated on the same day, notes were aggregated at the day level before evaluating the distribution of space savings. This aggregation reduces the influence of fragmented documentation patterns (e.g., multiple short addenda on the same date).

### 4.4 Clinical Evaluation of Summaries

We randomly selected 30 patients from the pilot set. A clinician (A.A.) evaluated all CLIN-SUMM summaries from these subjects at the note-level, individually scoring each summary from 698 notes, using a validation questionnaire. A second clinician (A.N.) independently assessed the summaries from 15 of these subjects again at the note-level, again individually scoring each summary from 403 notes.

The questionnaire comprised structured five-point scales (1=poor, 5=excellent) and yes/no questions to evaluate correctness, completeness, and hallucinations (**Supplementary Table 2**). Correctness verifies if the summary fully aligns with the key points in the notes without factual errors or misinterpretations. Completeness assesses the level of information coverage compared to the original notes and identifies any omissions. Finally, a yes/no question assesses whether the summary includes hallucinations or invented content not supported by the original notes. Although temporal coherence was not independently scored, errors in temporal fidelity directly impact the correctness and completeness scores as summaries are scored at the note level.

Inter-reviewer agreement was quantified using exact percentage agreement with 95% confidence intervals, unweighted Cohen’s κ, and quadratic weighted Cohen’s κ for ordinal ratings. Given the predominance of high scores and ambiguity in distinguishing adjacent rating categories, we additionally performed a sensitivity analysis dichotomizing scores into adequate (4-5) and inadequate (1-3). Positive and negative agreements were calculated for binary assessments. Confusion matrices were used to characterize the distribution and magnitude of disagreements.

To investigate whether errors accumulated during sequential summarization, each patient’s evaluated summary sequence was divided into early, middle, and late stages based on its relative position within that patient’s history. Mean correctness, completeness, and hallucination rates were compared across stages. Paired early-to-late comparisons were performed at the patient level using Wilcoxon signed-rank tests. We additionally investigated potential error propagation using a state-transition analysis of consecutive summaries. We estimated the probability of a hallucination in summary *i + 1* conditional on whether summary *i* contained a hallucination and calculated the corresponding risk difference and risk ratio.

### 4.5 Dementia Diagnosis and 3-year Prediction Models

#### 4.5.1 Data splits

We randomly stratified the cohort of 12,356 CLIN-SUMM summaries at the patient level into 2,612 training cases (21%), 802 validation cases (6.5%), and 604 test cases (5%), with an additional 8,338 cases (67.5%) reserved as an internal held out test set for further evaluation. The initial 4,018 subset was therefore used to derive and test the scaled model. For the final evaluation, results were reported on the combined test cohort (N = 8,942), consisting of the 604 internal test cases and the 8,338 internal held-out test cases (**Supplementary Table 9, 13**).

#### 4.5.2 Data, architecture and training

We trained models for two binary classification tasks: development of a dementia <u>diagnosis</u> model for dementia case identification from notes, and a dementia <u>3-year prediction</u> model for early clinical risk prediction. We fine-tuned the Clinical ModernBERT model ^22^, a transformer-based language model derived from BERT ^38^. BERT is a bidirectional transformer-based language model that learns contextual representations of text through self-attention. Clinical ModernBERT is an updated transformer architecture optimized for efficiency and longer context lengths and further pre-trained on clinical text to better capture medical terminology and documentation patterns. We trained the dementia diagnosis model using CLIN-SUMM summaries for all notes up to 30 days before the first formal dementia diagnosis (or for controls, 30 days before the censor date). As a sensitivity analysis, we repeated training and evaluation using notes available up to 90 days and six months before the first recorded diagnosis (**Supplementary Table 12**). We trained the dementia 3-year prediction model using CLIN-SUMM summaries for all notes up to 3 years before the first formal dementia diagnosis (or for controls, 3 years before the censor date). Notes were preprocessed by removing punctuation and special characters and the cleaned text was subsequently tokenized and converted to tensors. The Clinical ModernBERT model accepted sequences up to 8,192 tokens; for longer summaries, only the most recent summaries were used. To characterize the effect of this selection, we calculated the proportion of patients requiring truncation, the fraction of available tokens retained, the date of the oldest retained text, and retention of material dated more than five years before the diagnosis or matched reference date. We report these measures by case status and eligible note count in **Supplementary Tables 6–7**, with patient-level timelines in **Supplementary Figure 6**. Both the diagnosis and 3-year prediction models were trained using a binary cross-entropy loss with a learning rate of 1 × 10^−5^, batch size of 4, gradient accumulation of 4 and a weight decay of 0.05 for up to 8 epochs. A scheduler reduced the rate by 0.5 if validation loss plateaued for five epochs, and early stopping triggered after 2 stagnant epochs. The best-performing checkpoint was selected as the final model. For each task, we additionally trained Clinical ModernBERT on raw notes and trained TF-IDF ^23^ classifiers on both raw notes and CLIN-SUMM summaries. TF-IDF weights terms by their frequency within a document and their rarity across documents. Inputs were restricted to information available before the same task-specific cutoff. When text exceeded 8,192 tokens, we retained the most recent portion; TF-IDF used the same selected text as Clinical ModernBERT for each representation. Further details are provided in **Supplementary Section 3 and Supplementary Table 8**.

#### 4.5.3 Age-sex baseline

As a reference model, we trained a logistic regression classifier using only age and sex as predictors. For the diagnosis and 3-year prediction models, age was defined as the patient’s age 30 days or 3 years prior to the diagnosis/censor date, respectively. Age was standardized prior to model training. Because the difference between 30 days and 3 years represents a relatively small-time shift, the standardized age values, and therefore the resulting model performance metrics, were nearly identical between the diagnosis and 3-year prediction models. This baseline therefore provides a minimal demographic benchmark for comparison with models incorporating additional clinical information.

#### 4.5.4 Age-sex-comorbidities baseline

We next trained a logistic regression baseline incorporating age, sex, and prevalent comorbidities derived from ICD and SNOMED coded diagnoses. Comorbidities included anemia, arthritis, depression, delirium, osteoporosis, dizziness, syncope, hyperlipidemia, dyspnea, Parkinson’s disease, hearing impairment, visual impairment, anorexia, anxiety, alcohol use disorder, hypertension, and history of falls (**Supplementary Table 18**). The selected conditions were based on established dementia risk factors and clinical variables included in previously published dementia risk scores, including the CAIDE ^25^ dementia risk model. Each condition was represented as a binary indicator denoting the presence of a corresponding condition code 30 days or 3 years prior to the diagnosis/censor date, respectively.

#### 4.5.5 Age-sex-comorbidities-vitals baseline

To assess whether routinely collected physiologic measurements improve predictive performance, we extended the age-sex-comorbidities baseline to include vital signs and lab values commonly used in dementia risk models, including those included in the CAIDE dementia risk score.

Specifically, we incorporated body mass index (BMI), total cholesterol, systolic blood pressure and diastolic blood pressure (**Supplementary Table 19**). For the diagnostic task, measurements were obtained from records within the period spanning 5 years to 30 days prior to the diagnosis/censor date, with the most recent available value used for each variable. For the 3-year prediction task, measurements were obtained from the period spanning 8 years to 3 years prior to the diagnosis/censor date, again using the most recent available value. Because these variables were not available for all individuals, inclusion of vital signs and lab values resulted in a reduced sample size due to missing data.

#### 4.5.6 Utilization baseline

To assess the contribution of documentation volume, we trained a utilization-only baseline using the number of eligible notes, years of available history, and available summary token count, all measured before the task-specific cutoff. We also evaluated these variables alongside age and sex, and alongside age, sex, and comorbidities. Models were evaluated on the same task-specific internal held-out test sets as Clinical ModernBERT (**Supplementary Table 11**). We summarized eligible note counts, available tokens, observation time, and the proportion of summary inputs exceeding 8,192 tokens by case status and task (**Supplementary Table 10**).

#### 4.5.7 Fusion model

For both the diagnosis and three-year prediction tasks, we trained a logistic regression fusion model combining Clinical ModernBERT prediction scores from CLIN-SUMM summaries with age, sex, comorbidities, vital signs, and healthcare utilization variables (**Methods Section 4.5.6**). All inputs were derived from information available before the task-specific cutoff. Because vital-sign data were unavailable for some patients, we evaluated the fusion model in the subset with the required inputs and compared it with the CLIN-SUMM summary only model on those same patients.

#### 4.5.8 Interpretability

To interpret the predictions of the fine-tuned Clinical ModernBERT dementia classification model, we applied Local Interpretable Model-agnostic Explanations (LIME) ^26^ to quantify the contribution of individual words toward model predictions. For each patient summary, LIME generates a set of perturbed versions of the original text by randomly masking subsets of words. The fine-tuned Clinical ModernBERT model is evaluated on each perturbed version to obtain predicted probabilities for the dementia class. LIME then fits a locally weighted linear surrogate model that approximates the behavior of the neural network around the original document.

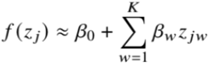

where zј represents a perturbed document expressed as a binary word-presence vector, zјw = 1 indicates that word *w* is present in perturbation *j*, and *βw* represents the local attribution coefficient for word *w*. Perturbations closer to the original document receive higher weights using an exponential kernel based on cosine distance, ensuring the surrogate model approximates the neural network locally.

For each patient we retained the top 100 words selected by LIME according to their contribution magnitude. These coefficients quantify the direction and strength of each word’s contribution to the model prediction. Positive coefficients indicate words that increase the predicted probability of dementia (risk-promoting words), while negative coefficients indicate words that decrease the predicted probability and support the control class (protective words). Because LIME perturbs words while Clinical ModernBERT processes subword tokens, these coefficients should not be interpreted as attributions to the model’s native tokens. We used this analysis to explore language associated with predictions in the selected patients.

To identify consistent linguistic patterns across individuals, word-level attribution coefficients were aggregated across patients. Interpretability analysis was conducted on 100 test cases, consisting of 50 high-risk individuals, randomly selected from the top 5% of predicted dementia risk (≥ 95^th^ percentile), and 50 low-risk individuals, randomly selected from the bottom 5% of predicted risk (≤ 5^th^ percentile). Selection was stratified by age and sex to ensure comparable demographic distributions between groups. For each word w, we computed the conditional attribution score, representing the average attribution coefficient across patients in whose LIME explanation the word was identified as an informative feature:

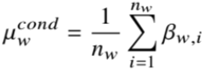

where *nw* denotes the number of patients in whose explanation word *w* appears, and *βw, i* represents the attribution coefficient of word *w* for patient *i*.

Word attribution results were visualized using a category-structured text plot in which words were arranged by semantic category and positioned along a horizontal axis according to their attribution score. Words positioned to the right of zero represent terms that increase the predicted probability of dementia (promoting words). Words positioned to the left of zero represent terms associated with the healthy class (protective words). Font size scales with the magnitude of the attribution coefficient, allowing visually prominent representation of strongly influential words. Words were grouped into predefined categories (Neurocognitive, Symptoms/Signs, Conditions/Diagnoses, Drugs/Medications, Clinical Documentation, Clinical Procedures/Tests, Descriptors/Qualifiers, Demographics, Body Parts/Anatomy, Lifestyle/General Health, Other, Dosage/Administration, Units/Measurements), with distinct colors assigned to each category. Intensity of color represents the prevalence of the word across the cohort. Within each category, words were ranked by the absolute value of their attribution score, and the 10 top-ranked terms within each category were displayed for both positive and negative contributions. Categories such as Other, Dosage/Administration and Units/Measurements were excluded from the final visualization because the associated terms were largely non-informative and did not meaningfully contribute to model interpretation.

As a tokenization-aware sensitivity analysis, we applied Integrated Gradients (IG) ^29^ to the original, unperturbed inputs for the same patients. IG estimated signed attributions for Clinical ModernBERT’s native subword tokens by integrating gradients of the dementia-class score from a reference embedding to the input embedding. We summed attributions across embedding dimensions and then across subwords belonging to the same word occurrence. Scores for repeated occurrences were averaged within patients and then across patients containing the word. We compared IG with the features already selected for Figure 5; IG was not used to select additional features. We assessed agreement in attribution direction and rank correlation between the two methods (**Supplementary Figure 8**).

### 4.6 Medication Extraction

Medication usage events were extracted from the CLIN-SUMM patient summaries. Each patient summary contains a dedicated section describing medication history aggregated across clinical notes. Using these condensed summaries enables efficient downstream processing, as CLIN-SUMM summaries provide a compact representation of clinically relevant information while preserving temporal context across visits.

Medication extraction was performed using GPT-4o. This was a separate downstream analysis: Qwen3 generated the longitudinal representation, while GPT-4o converted medication mentions in that representation into dated, structured events. We did not evaluate Qwen3 for medication extraction. For each patient summary, the model was prompted to identify medication usage events for a predefined set of dementia-related and neuropsychiatric medications relevant to our cohort: Donepezil, Galantamine, Memantine, Rivastigmine, Lecanemab, Donanemab, Suvorexant, and Brexpiprazole. Brand and generic names were treated as equivalent. The model was instructed to extract all medication events mentioned in the summary and return them in structured JSON format, including the event date (ISO format, when available) and the current dose in milligrams if specified. If a medication was discontinued, the dose was recorded as 0 mg on the corresponding date.

The following prompt was used:

> *Given the following summary of patient notes, extract all mentioned medication usage events for Donepezil, Galantamine, Memantine, Rivastigmine, Lecanemab, Donanemab, Suvorexant, and Brexpiprazole. Treat brand and generic names as equivalent. For each medication, return a list of events including the date (YYYY-MM-DD if possible) and the current dose in mg if mentioned. If discontinued on a particular date, assign 0 mg to the current dose. Return the result strictly as valid JSON using the structure {“MED 1“:[{“date“:“YYYY-MM-DD”,“dose_mg“: <value>}], … }*.

Using medication events extracted from CLIN-SUMM summaries, we reconstructed longitudinal medication trajectories for dementia-related treatments. To validate this approach and evaluate its ability to recover medication information not captured in structured EHR data, we compared LLM-extracted medication events from CLIN-SUMM summaries with medication records obtained from Snowflake tables. Structured records were available from 2015 onward, whereas summaries could contain events from earlier notes. Structured records could also extend beyond the last note included in a summary. We therefore restricted the direct comparison to each patient’s shared observation window, beginning in 2015 and ending on the earlier of the last included note or the study censoring date. Trajectories in Figure 6B were constructed from the full set of eligible CLIN-SUMM-extracted events and were not restricted to this shared window.

To evaluate dose concordance, we selected one CLIN-SUMM event and structured record pair per patient, initially choosing the pair with the smallest difference between the summary event date and structured medication order date. When multiple strengths were recorded on the same or nearby dates, we allowed an alternative pair with matching doses if its date gap was no more than two days greater than the minimum. We assessed exact dose agreement for the selected pairs.

### 4.7 Evaluation of a Commercial Frontier LLM

To evaluate the CLIN-SUMM framework across different model families, we implemented the pipeline using a commercial frontier model (GPT-4o) accessed via the OpenAI API in the pilot subset. This evaluation was designed to provide a reference point for summarization quality and to enable comparison with an open-source alternative (Qwen3) used for large-scale deployment. Because frontier models differ in context capacity, instruction following, and API constraints, several implementation adjustments were made. GPT-4o required fewer prompt constraints to achieve structured output. For example, explicit enumeration of laboratory test names and detailed examples used in the Qwen prompt were no longer necessary. The pipeline incorporated a sliding context window to ensure that longitudinal summaries remained within the model’s input limits. For GPT-4o, a window size of 50 prior summaries was used, representing the largest configuration that maintained stable API performance while avoiding request size limits and latency constraints. Specifically, it avoids intermittent rate-limit and timeout errors that stemmed not from token overflows but from API-level constraints, including transient throughput throttling, payload-size serialization overhead, and server-side latency caps on long requests, leading to slower response times and occasional failed generations. Overall, the frontier model produced stable and well-structured summaries with minimal prompt modification.

The GPT-4o evaluation was conducted on the pilot dataset to allow direct comparison with summaries generated by Qwen3 under identical conditions. To assess textual similarity between summaries generated by GPT-4o and Qwen3, we computed BERTScore ^39^, which measures semantic similarity between two texts using contextual embeddings, with GPT-4o output serving as the comparison text; neither output was treated as a source-note-verified reference standard. BERTScore evaluates token-level alignment between candidate and reference texts in embedding space and reports precision, recall, and F1 scores. Precision measures how well tokens in the candidate summary match semantically relevant tokens in the reference summary, recall measures how well the candidate summary captures the semantic content of the reference summary, and the F1 score provides a balanced measure of both coverage and fidelity. These metrics were computed between GPT-4o-generated summaries and Qwen3-generated summaries for the same patient records.

## 5 Data availability

The Massachusetts General Brigham source data are not publicly available because they are electronic health records. Making the data publicly available without additional consent or ethics could compromise privacy.

## 6 Code availability

The LLM prompts are provided in their entirety in **Supplementary Figure 1**. Code for the sliding window, Jaccard similarity evaluation and Clinical ModernBERT fine-tuning for disease diagnosis and 3-year prediction will be made publicly available via a public repository upon publication.

## 7 Acknowledgments

M.M. is partially supported by grants from the National Institutes of Health (3OT2OD035404-01S3, 1R01NS134597, and 1UG3HG014379-01) and from the American Heart Association (961045). C.D.A. reports funding by NIH U01NS069673, RF1NS139183, R01NS093870, AHA 21SFRN812095, and the Global Brain Care Coalition.

## 8 Author contributions

V.D. contributed to methodology, data curation, formal analysis, visualization, and writing. D.F.P. contributed to methodology, visualization, and writing. A.A., A.N., and E.B.H. contributed to clinical validation and investigation. W.H. contributed to data curation. T.N. and S.F. contributed to methodology. S.J.A. contributed to data curation, clinical conceptualization and interpretation. C.D.A. contributed to clinical conceptualization and interpretation. J.H. contributed to conceptualization and funding acquisition. M.M. contributed to conceptualization, methodology, funding acquisition, and supervision. All authors contributed to review and editing of the final manuscript.

## 10 Human Ethics and Consent to Participate

This study protocol was approved by the Institutional Review Board (IRB) of Massachusetts General Brigham (#2023P001077) and complied with the Declaration of Helsinki. The requirement for informed consent was waived by the Institutional Review Board given the low-risk retrospective nature of the study and privacy-protecting measures implemented by the investigators.

