## Supplemental for "CLIN-SUMM: Incremental Longitudinal Summarization of Clinical Notes Enables Scalable Representation and Early Disease Prediction"

**Supplementary Material**

**Index**

**Section 1:** Prompt Optimization and Parameter Selection

**Section 2:** Error Analysis of CLIN-SUMM Summaries

**Section 3:** Comparison with Raw Clinical Notes

**Section 4:** Early detection time horizon

**Section 5:** Additional analyses of model behavior

**Section 6:** Additional Applications of CLIN-SUMM Summaries

**Supplementary Figure 1:** Two-step summarization

**Supplementary Figure 2:** Compression increases with the number of processed notes

**Supplementary Figure 3:** Distribution of average summarization space savings per individual

**Supplementary Figure 4:** Inter-rater agreement on summary quality

**Supplementary Figure 5:** Summary accuracy and potential error propagation

**Supplementary Figure 6:** Clinical history retained at each task cutoff

**Supplementary Figure 7:** Mean predicted dementia risk across age and time relative to the diagnostic endpoint

**Supplementary Figure 8:** Agreement between LIME and Integrated Gradients attributions

**Supplementary Figure 9:** Mean predicted dementia risk across patient subgroups under different summary truncation conditions

**Supplementary Table 1:** Demographic characteristics for the dementia, control and entire study cohorts

**Supplementary Table 2:** Structured summary evaluation questionnaire

**Supplementary Table 3:** Validation results for CLIN-SUMM summaries

**Supplementary Table 4:** Inter-rater agreement for clinical summary assessments

**Supplementary Table 5:** Summary quality across patient histories and hallucination transitions

**Supplementary Table 6:** Retained history for the Diagnosis model (30-day cutoff)

**Supplementary Table 7:** Retained history for the 3-year Prediction model

**Supplementary Table 8:** Performance of summary and raw-note representations

**Supplementary Table 9:** Performance of the Clinical-ModernBERT models in the test cohort using CLIN-SUMM summaries for dementia diagnosis and 3-year prediction, compared to baseline logistic regression models

**Supplementary Table 10:** Documentation volume and history availability by case status and task

**Supplementary Table 11:** Performance of utilization-based models and Clinical ModernBERT on CLIN-SUMM summaries

**Supplementary Table 12:** Sensitivity of Clinical ModernBERT performance to the cutoff before the first recorded dementia diagnosis

**Supplementary Table 13:** Time-horizon evaluation using the Diagnosis model applied to earlier cutoffs

**Supplementary Table 14**: BERTScore comparison between Qwen3- and GPT-4o-generated CLIN-SUMM summaries

**Supplementary Table 15:** ICD-9 and ICD-10 diagnostic codes used to ascertain dementia

**Supplementary Table 16:** SNOMED diagnostic codes used to ascertain dementia

**Supplementary Table 17:** Performance metrics by distance bin, including similarity, recall, and information loss, used to determine similarity threshold for Jaccard distance

**Supplementary Table 18:** Diagnostic codes used to extract comorbidities from the EHR

**Supplementary Table 19:** SNOMED codes (measurement table) used to extract vital signs and laboratory values

### 1 Prompt Optimization and Parameter Selection

CLIN-SUMM uses two distinct prompts to iteratively summarize longitudinal clinical notes (**Supplemental Figure 1**). We refined the two prompts following best practices 1, evaluating several variants for each component. The chosen approach involves guiding the model to produce structured responses and defining the desired behavior when no new content was identified, improving performance over autonomous generation.

While the CLIN-SUMM framework is model-agnostic and compatible with any prompt-based LLM API, we used the Qwen3 model 2, specifically the 14B parameter variant (`Qwen/Qwen3-14B` checkpoint) [3], deployed through the vLLM inference framework (v0.12.0) [4], on an Azure Standard NC24ads A100 v4 virtual machine equipped with one NVIDIA A100 GPU, 24 virtual CPUs, and 220 GiB of system memory. We did not apply quantization. To achieve good results using this LLM, we applied several prompt refinements and adjustments: (1) we disabled “think mode” and enforced stricter structural guardrails to ensure consistent sectioning; (2) we provided concrete examples of commonly missed clinical elements, such as specific laboratory names (e.g., TSH, HbA1c, CBC, BMP) and vital signs to improve coverage of quantitative clinical details; and (3) we increased top-p and top-k values to 0.9 and 60, respectively, to encourage the model to retain more of the clinical terms that open-source models tend to truncate under default decoding settings.

To manage longitudinal context while respecting model constraints, CLIN-SUMM integrates a sliding window mechanism in the second summarization prompt. This window processes only the most recent summaries rather than the complete clinical history, allowing the model to identify new information while maintaining continuity across encounters. For the Qwen implementation, the sliding window size was set to 25 summaries to ensure stable inference, reflecting a practical limitation imposed by the vLLM framework’s decoder prompt cutoff. In preliminary experiments, contexts exceeding approximately 25 summaries frequently resulted in generation failures due to prompt-length constraints. A dynamic safeguard was also implemented to handle edge cases in which the 25 summaries exceeded the allowable token limit, leading to runtime failures. In these cases, the system automatically reduces the number of summaries included in the prompt to remain within the decoder limit. Such adjustments occurred infrequently and affected only a small fraction of encounters. In the final-run logs, 4,170 updates (0.15%) used a reduced history window, and 251 notes (0.009%) required chunking. We identified no unhandled context-length errors in the completed runs, but development-stage errors were not systematically counted and the final logs were not a complete request-level failure audit. We therefore cannot estimate an overall generation-failure rate reliably. In addition to technical stability, the fixed window size of 25 was chosen to ensure methodological consistency across patients. Because note volume varies widely by patient (some with fewer than 20 encounters, others exceeding 100), using a standardized context size avoids biasing the downstream machine learning tasks towards patients with longer documentation histories. Finally, fixing the sliding window size facilitates seamless adaptation to alternative LLMs with increased reproducibility across implementation, if desired.

CLIN-SUMM implements a Jaccard similarity evaluation [5] between consecutive notes. If this similarity is high, the subsequent note is skipped to avoid redundant summarization of copy-pasted content, conserving tokens and API request charges. A conservative threshold of 95% was set to retain subtle new information, which we chose empirically by computing entity-level recall (i.e., proportion of retained clinical entities) between consecutive notes as a function of Jaccard distance (**Supplemental Table 17**). We observed that entity recall remained above 94% for note pairs with Jaccard distance ≤ 0.05 (≈ 95% similarity), but dropped sharply beyond that range. This inflection point indicates that notes exceeding this distance begin to introduce new clinical entities or contextual updates rather than templated repetition. Accordingly, we set a 0.05 distance threshold (95% similarity) to discard near-duplicate notes while preserving semantically distinct updates, minimizing information loss to < 6% on average.

We also explored the LLMs temperature hyperparameter, which influences its conditional probability distributions during sampling and thus affects the likelihood of less probable token generation. Higher temperatures increase randomness and “creativity,” while lower temperatures yield more deterministic and faithful outputs. We empirically determined that a temperature of 0.2 provided the best qualitative balance between completeness and factual precision, generating concise and stable summaries while maintaining high fidelity to the source notes. In exploratory runs, higher values (> 0.2) produced summaries that were longer but less stable, occasionally introducing mild factual drift (e.g., adding diagnosis from clinical conditions not explicitly mentioned in the source notes). Conversely, lower values (< 0.1) yielded overly compressed outputs that omitted secondary but clinically relevant information such as medication adjustments or lab trends.

A measured batch of 2,914 notes took 11,248 seconds to summarize, averaging 3.86 seconds per note. At the Azure virtual-machine rate used in our estimate ($3.30 per hour), extrapolation to an illustrative workload of 100 patients with 100 notes each gives approximately $35 in compute charges. This is not a fixed per-note cost: later updates can include more prior summary text, and runtime varies with note length, patient history, and batching. The approximately $250 GPT API estimate depends on input and output token volumes and the API prices applicable at the time of use.

### 2 Error Analysis of CLIN-SUMM Summaries

Some examples of LLM model errors were: (1) a clinical note mentioning “increasing back pain, dysuria, discolored urine, advised patient to go to [urgent care]” summarized as “Diagnosis: . . . possible urinary tract infection or other renal condition” (although the model may be correct, it is interpolating a diagnosis when none is mentioned in the original note), (2) a primary care note with “HCM: colonoscopy was done in . . . ” mistakenly detected as a diagnosis of “hypertrophic cardiomyopathy” instead of ‘healthcare maintenance”, and (3) completeness errors where the summary does not include patient adherence to therapy or pertinent negatives, such as “no AFIB detected on the ziopatch”.

### 3 Comparison with Raw Clinical Notes

We compared CLIN-SUMM summaries with raw clinical notes for both classification (30 days before the first recorded dementia diagnosis) and 3-year prediction task. Separate Clinical ModernBERT models were trained for each representation and task. Inputs were limited to 8,192 tokens; when more text was available, the most recent text was retained. For each task and representation, a TF-IDF [6] model was trained on the same selected text used for Clinical ModernBERT. Models were evaluated on the same internal held-out patients for their respective tasks.

Within the fixed input budget, selected raw notes represented approximately 0.7 years of documentation, whereas summaries represented nearly 2 years. The comparison therefore evaluates CLIN-SUMM's complete representation, including its greater historical coverage within the input limit (**Supplementary Table 8)**.

**4 Early detection time horizon**

- **Longitudinal AUC figure:** To evaluate how model performance varies as a function of prediction horizon and to assess the risk stratification capability of the dementia classification model, we conducted longitudinal analyses across multiple time windows preceding the diagnosis date (**Figure 4B**). At each time horizon *X* predictions were generated by applying the dementia classification model using only clinical information available up to *X* years before the dementia diagnosis date (or censoring date for controls). This setup allowed us to evaluate how effectively the model stratifies individuals by dementia risk when applied at progressively earlier points in the clinical timeline. Model performance was evaluated across time windows extending up to 5 years prior to the index date. Performance improved progressively as the prediction window approached the diagnosis date, reflecting the increasing availability of clinically relevant signals in the record. Notably, the model retained sufficient discriminatory power several years prior to diagnosis, indicating its ability to stratify individuals by risk earlier in the disease trajectory. The analysis was limited to a 5-year horizon because data availability and cohort size decreased substantially beyond this point, leading to unstable estimates.

- **Longitudinal risk trend lines:** To characterize how model predictions from the dementia classification model evolve over time prior to diagnosis, we constructed longitudinal prediction trajectories using cumulative yearly windows of patient summaries. As illustrated in **Figure 4C**, predictions were generated at one-year increments, where each step incorporated all clinical information available up to that time point. At each step K, the input to the dementia prediction model consisted of the cumulative set of summaries from the beginning of the patient record up to K years before the endpoint. This cumulative yearly evaluation was designed to mimic how risk might be reassessed during annual clinical visits. The procedure produced a sequence of predicted dementia probabilities for each patient across progressively earlier time windows. Because patients have varying lengths of clinical history, trajectories were right-aligned relative to the endpoint (diagnosis for cases and censoring for controls). For each yearly window, we computed the mean predicted probability across patients within each group (dementia vs. control). To ensure stable estimates, only windows with at least 50 individuals in the dementia cohort were included in the analysis. The upper panel of **Figure 4C**, shows the mean predicted dementia risk over time for both groups, with linear trend lines indicating overall trajectory direction. The middle panel visualizes the distribution of predicted probabilities at each time window using kernel-density ridge plots (violin-style distributions). The lower panel reports the number of individuals contributing predictions at each window. An interval spanning approximately 3–6 years before diagnosis was highlighted as a region where divergence between dementia and control trajectories becomes most pronounced.

- **Longitudinal risk with time bins:** To further examine how predicted dementia risk varies jointly as a function of age and time before diagnosis, we constructed a two-dimensional risk surface using the predictions generated by the dementia classification model. For each patient, the model produced a sequence of predicted probabilities across cumulative yearly windows prior to the endpoint (diagnosis for cases and censoring for controls). The final window corresponds to the endpoint ($K$), with earlier windows representing progressively earlier time points ($K-1$, $K-2$, . . .). To isolate the effect of age at each time window, we estimated the patient's age at each window by back-calculating from the age recorded at the endpoint. Predictions were then stratified jointly by age bin and time window bin ($K$). Within each (age bin, $K$) cell, we computed the mean predicted dementia probability separately for dementia cases and controls. To ensure stable estimates, cells containing fewer than 10 individuals were excluded from the analysis. Additionally, the maximum time window included was limited to the longest window supported by the dementia cohort to ensure comparable support between groups. The resulting age–time grid was visualized as a three-dimensional surface showing the mean predicted dementia risk across age and time relative to the endpoint (**Supplementary Figure 7**). Contour projections were also included to highlight gradients in predicted risk. To further illustrate disease-specific patterns, we computed a difference surface by subtracting the control risk surface from the dementia surface, highlighting regions of the age–time space where the model predictions diverge most strongly between groups. This analysis provides a joint view of how predicted dementia risk evolves over time while accounting for the changing age of individuals across longitudinal observation windows.

### 5 Additional analyses of model behavior

#### 5.1 Misclassifications

To better understand model errors and identify systematic patterns in misclassification, we performed a post hoc analysis of the representation space learned by the model. For both the training and validation datasets, we obtained vector representations (embeddings) generated from the classification model and identified prototypical control and dementia reference points using highly confident correctly classified individuals in the training set. True control cases were defined as patients with true label 0 and predicted label 0 whose predicted probability of dementia fell within the lowest decile of model predictions. Conversely, true dementia cases were defined as patients with true label 1 and predicted label 1 whose predicted probability of dementia fell within the highest decile of model predictions. For each group, we computed the centroid of the embedding vectors by averaging the embedding coordinates across the selected individuals, yielding representative control and dementia reference embeddings. For each individual, we then computed the Euclidean distance between the individual's embedding vector and each centroid. A relative similarity score was defined as:

$$S_i = d(\mathbf{x}_i, C_{\text{ctrl}}) - d(\mathbf{x}_i, C_{\text{dem}})$$

where $d(\cdot)$ denotes the Euclidean distance, $\mathbf{x}_i$ represents the embedding vector for individual $i$, and $C_{\text{ctrl}}$ and $C_{\text{dem}}$ denote the centroid embeddings of the reference control and dementia groups, respectively. Positive values of $S_i$ indicate that an individual lies closer to the dementia centroid than to the control centroid in embedding space, whereas negative values indicate greater similarity to the control centroid.

We then identified two high-confidence discordant groups by combining the embedding similarity metric with model prediction probabilities. First, control-like dementia cases were defined as individuals with true dementia labels whose embeddings were closer to the control centroid and whose predicted dementia probability fell within the lowest decile of predictions (*n* = 11 in training set, *n* = 8 in validation set). Second, dementia-like control cases were defined as individuals with true control labels whose embeddings were closer to the dementia centroid and whose predicted dementia probability fell within the highest decile (*n* = 13 in training set, *n* = 2 in validation set). These groups represent highly confident misclassifications in which both the learned representation and the model prediction align in suggesting the incorrect class. Cases within these subsets were prioritized for manual review to characterize systematic sources of model error.

We manually reviewed approximately half of these cases in detail (*n* = 21), revealing several recurring failure modes. False positives (controls misclassified as dementia cases) most commonly occurred in patients with cognitive complaints attributable to non-degenerative conditions, including major depressive disorder, prior structural brain injury, or other neurologic comorbidities, where documentation contained dense cognitive vocabulary (e.g., “memory loss,” “neuropsychological testing,” “cognitive impairment”) despite preserved functional status and negative formal testing. In some cases, age- and vascular risk–related language (e.g., stroke family history, hearing loss, hypothyroidism, advance directives) may have contributed to overestimation of dementia probability in otherwise cognitively intact individuals. Notably, the study was conducted using data available through 2023; however, a post hoc analysis using subsequently available data through 2025 identified dementia diagnoses in 2 of 15 false positive cases. This suggests that some apparent false positives may reflect delayed clinical diagnosis or prodromal disease that was too early in its course to be diagnosed within the original study period. False negatives (dementia cases misclassified as controls) were observed in two principal contexts: (1) patients whose actual dementia symptoms and diagnosis occurred after the study’s data cutoff; and (2) patients with sparse cognitive documentation, particularly when care was concentrated in specialty clinics (e.g., dermatology, urology, procedural services) where cognitive status was minimally assessed or recorded. Additionally, early or mild presentations without clear documentation of functional decline appeared more likely to be under-detected.

Overall, these findings suggest that most misclassifications arise from ambiguous or incomplete clinical context rather than fundamental model limitations. In particular, errors often reflect cases where clinical language is suggestive but non-specific, especially cognitively dense language in controls, or where documentation is sparse across encounters,  or delayed diagnostic labeling in real-world records.

#### 5.2 Model Predictions Relative to Mild Cognitive Impairment Diagnosis

To assess how the model behaves in the presence of early cognitive impairment signals, and to better understand factors contributing to borderline predicted risk among controls, we conducted a subset analysis based on diagnostic codes associated with mild cognitive impairment (MCI), including G31.84 (Mild Cognitive Impairment), 331.83 (Mild cognitive impairment, so stated), R41.81 (Age-related cognitive decline), and R41.840 (Attention and concentration deficit). We identified individuals with at least one MCI-related code and categorized them based on whether the code appeared before diagnosis/censoring, after diagnosis, or not at all.

Across the CLIN-SUMM cohort (n = 6,178 per group), 2,092 dementia patients (33.9%) and 212 controls (3.4%) had at least one MCI-related code. Among dementia patients with an MCI code, 911 (14.8%) had the code recorded before diagnosis, while 1,181 (19.1%) had it recorded after diagnosis. In the control group with an MCI code, 82 (1.3%) had the code recorded prior to censoring, while 130 (2.1%) had it recorded after the censoring date, while 5,966 controls (96.6%) had no recorded codes.

Within the test set, 608 dementia patients (13.6%) had an MCI code before diagnosis and 854 (19.1%) after diagnosis, while 3,009 (67.3%) had no MCI-related code recorded. Among controls, 53 individuals (1.2%) had an MCI code before censoring, 89 (2%) had after censoring and 4,329 (96.8%) had none. To assess how early cognitive impairment signals influence model predictions, we evaluated predictions from the diagnosis model within these groups in the test set. In an additional analysis, summaries were truncated at the time of the first MCI code, allowing us to assess whether the clinical information available up to the point when MCI is first recorded already contains sufficient signals for predicting dementia risk.

Mean predicted dementia probabilities across these groups are shown in **Supplementary Figure 9.** Dementia patients with MCI codes recorded prior to diagnosis had the highest predicted probabilities, while controls without MCI codes had the lowest. Controls with an MCI code showed intermediate predictions and were notably closer to the dementia group in the predicted probability distribution. When summaries were truncated at the time of the first MCI code, predicted probabilities decreased across groups, suggesting that additional clinical signals appearing later in the record contribute to the model's predictions.

### 6 Additional Applications of CLIN-SUMM Summaries

#### 6.1 Disease Progression Analysis

We performed an exploratory assessment of whether **CLIN-SUMM** summaries could support automated analysis of disease trajectories. We provided the GPT-4o large language model (LLM) with patient-level **CLIN-SUMM** summaries and prompted it to extract a structured view of "Condition Evolution & Disease Progression". For a representative patient, the model produced the following output:

1. **Initial Focus (2017–2018):** The early records highlight generalized anxiety disorder, sleep apnea, and gastroesophageal reflux disease (GERD). Symptoms include anxiety attacks,snoring, and sleep disturbances. During this time, medication adjustments (e.g., Sertraline to Venlafaxine) were frequent due to side effects like sexual dysfunction.

2. **Mid-course (2018–2019):** There's clear progression with the emergence of chronic cough, back pain, and vertebral compression fractures (due to coughing or osteopenia). This time frame also marks hospitalizations for pericardial effusion and early signs of cardiac disease (e.g., chest pain, syncope).

3. **Advanced Disease (2020–2023):** The patient experienced a STEMI (myocardial infarction) with stent placement and later AV block, necessitating a pacemaker. There's evidence of ischemic cardiomyopathy, and recurrent angina despite prior interventions. Progressive osteoporosis, recurring facial flushing, and chronic sinusitis persist.

4. **Recent Status (2022–2023):** The patient deals with continued back pain, increased medication burden, and new symptoms such as vision changes, fatigue, and possible autonomic symptoms (e.g.,

facial flushing with elevated tryptase levels suggesting mast cell activation or similar disorders).

This breakdown reveals evolving patterns in diagnoses, medications, and comorbidities, demonstrating the ability of **CLIN-SUMM** summaries to reflect disease progression over time.

#### 6.2 Symptom Trend Tracking

We also explored whether longitudinal **CLIN-SUMM** summaries can reveal worsening, improved, or recurrent symptom trajectories. Again, we provided the GPT-4o LLM with patient-level **CLIN-SUMM** summaries and prompted it to extract a structured view of "Symptom Trends". In the same representative patient, the model produced the following output:

1. **Worsening: Chronic back pain** has become more debilitating, despite physical therapy and injections. **Facial flushing and syncope** have increased in frequency and complexity. Chest discomfort episodes are recurring, even **post-STEMI and PCI**.

2. **Improving or Stabilizing: Anxiety symptoms** appearto bemore controlled recently with escitalopram(Lexapro). **Sinus-related issues** are now part of a stable chronic management plan with allergy and ENT follow-ups.

3. **Recurring: GERD** and **postnasal drip, cough, and sleep apnea symptoms** have persisted throughout, with varying intensity.

Such trends illustrate the potential for **CLIN-SUMM** to support trajectory analyses of symptoms relevant to clinical trial eligibility, clinical monitoring and diagnosis.

### Supplementary Figures

#### (a) Prompt 1: Summarizing the first note

**Prompt template for summarizing first note of a patient**

```
Medical visit note (Timestamp: {note.name}):{note['text']}

Please generate a structured, comprehensive summary of the above medical note in an
abstractive manner, ensuring that the key details are captured and presented clearly and
redundant repetition is avoided

Organize the summary using the following sections:
**Chief Complaints**: [Summarize the primary concerns or symptoms reported by the patient]
**History of Present Illness**: [Provide a narrative of medical history for the symptoms
mentioned today, their onset, progression, associated factors, and any relevant context]
**Past Medical History**: [Summarize relevant past diagnoses, surgeries, or chronic
conditions]
"**Medications & Allergies**: [List current medications, dosages, and any known drug or food
allergies]
**Vital signs or Lab/Imaging Findings**: [Summarize vital sign measurements, any lab results
like hemoglobin and others, Physical Exam results or imaging results if any or respond
'nothing observed']
**Diagnosis**: [Summarize diagnoses made]
**Treatment Plan and Follow-up**: [Summarize the treatment plan including medications,
interventions, or lifestyle recommendations and outline if any follow-ups, referrals, or
additional tests were requested]

Ensure the summary is coherent, natural, and does not merely extract sentences from the text
but instead synthesizes the information into a well-structured report
```

#### (b) Prompt 2: Longitudinal update with changes

**Prompt template for summarizing subsequent notes of a patient**

```
Previous summary before (Timestamp: {current_note.name}):{previous_summary}
New visit note (Timestamp: {current_note.name}):{current_note['text']}

Identify any **new information** in the current visit that is not already included in the
past visit summaries
Update the relevant sections with new details while avoiding repetition
If a section has no new information, respond with 'nothing new'.

Format the updated summary with the following sections and definitions:
**Chief Complaints**:..................................
**History of Present Illness**: ........................................
.........................................
.........................................
**Diagnosis**: ............not mentioned before]"...........
**Treatment Plan and Follow-up**: .......................................
**Changes Over Time**: [Outline changes observed over time if any or respond 'nothing
observed'. Exclude: social history, family information, occupation, living situation, or other
non-medical details.]
**Ensure that the updated summary incorporates only new and relevant details while keeping
the structure consistent...
```

**Supplementary Figure 1: Two-step summarization:** (a) Initial note summary; (b) Incremental update with summarization of novel content and “changes over time”.

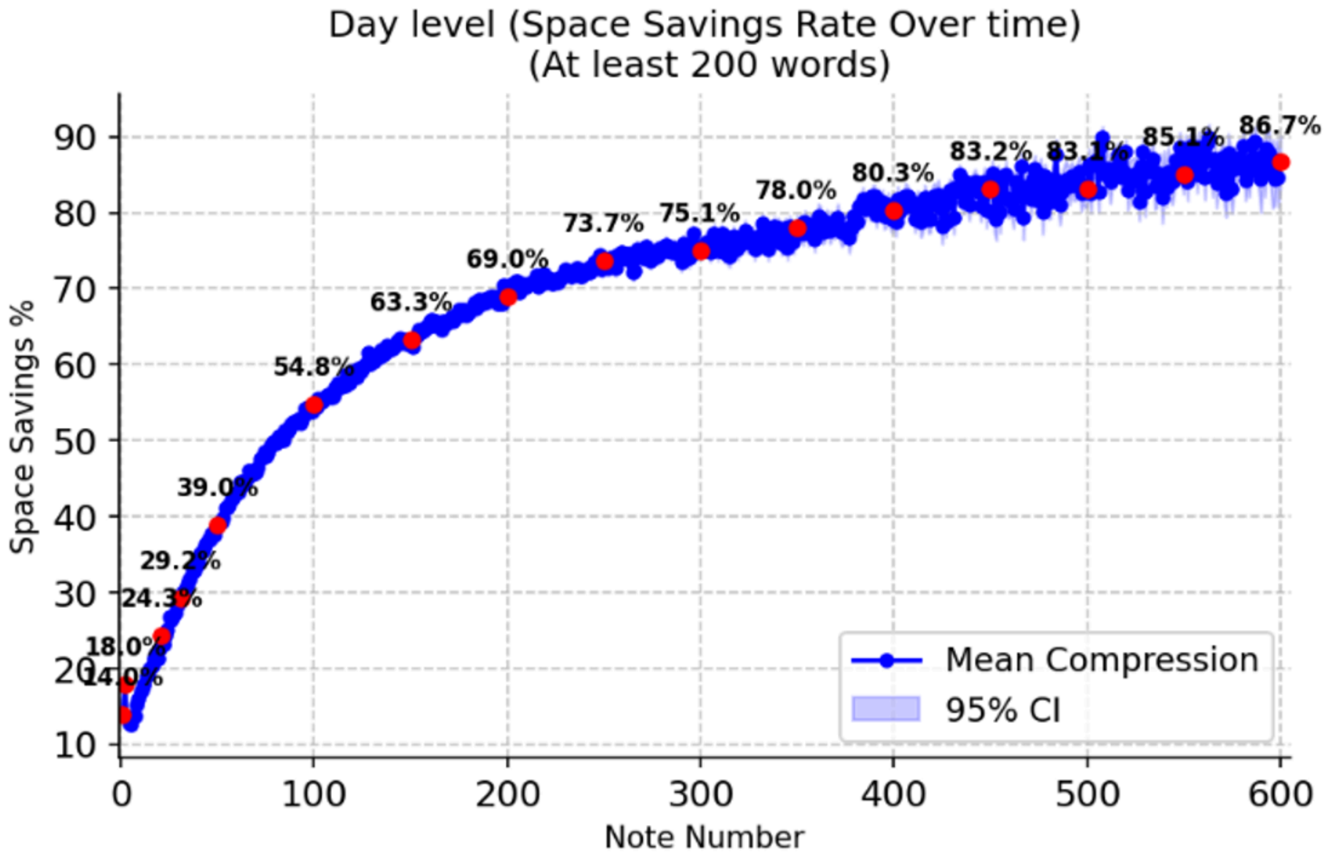


**Supplementary Figure 2:** On average, compression increases with the number of processed notes in the full cohort. The blue line represents the mean space savings achieved by CLIN-SUMM, the blue shaded region indicates the 95% confidence interval across patients, and red dots highlight key representative points. Larger confidence intervals on the right result from fewer patients with many notes.

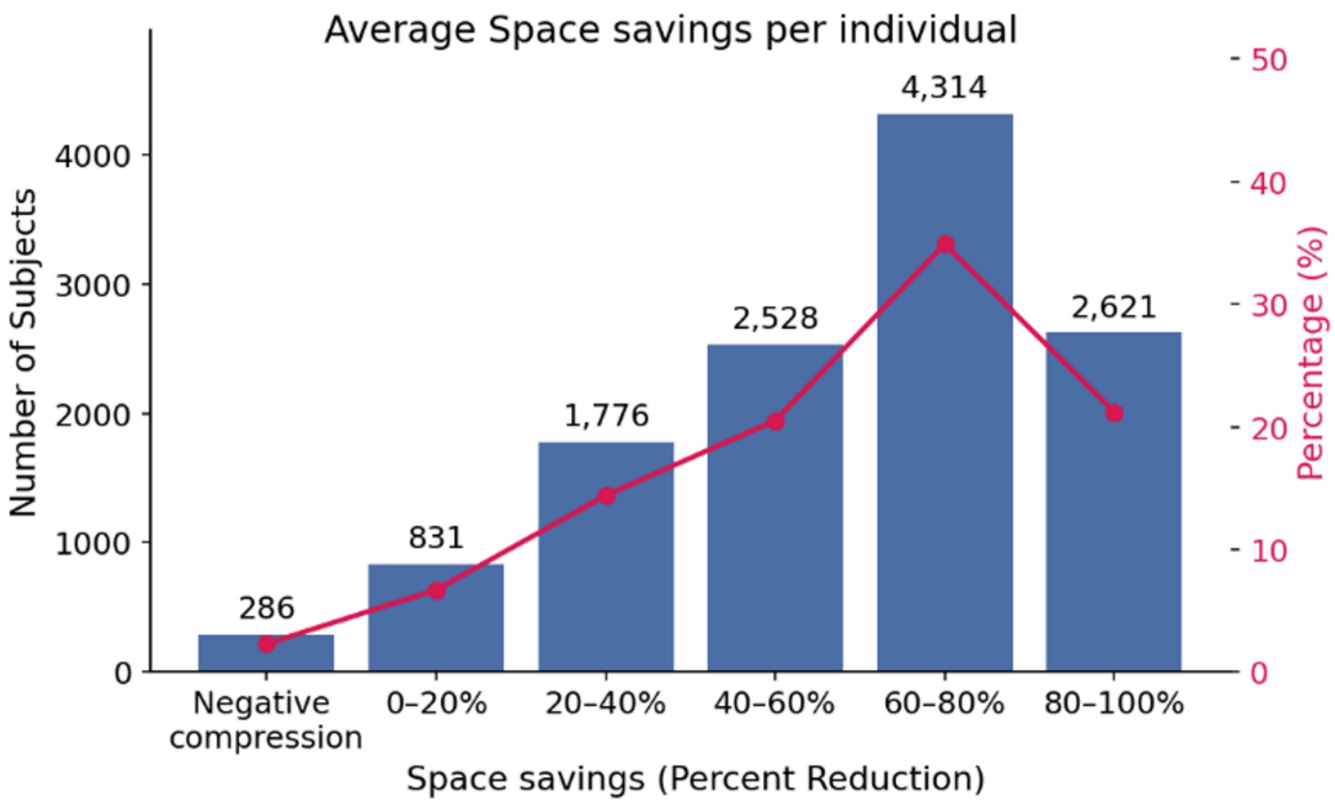


**Supplementary Figure 3:** Distribution of average summarization space savings per individual in the full cohort. Most individuals exhibit positive compression, with a very small subset showing negative compression, largely driven by short notes or less informative note types (e.g., return or phone notes).

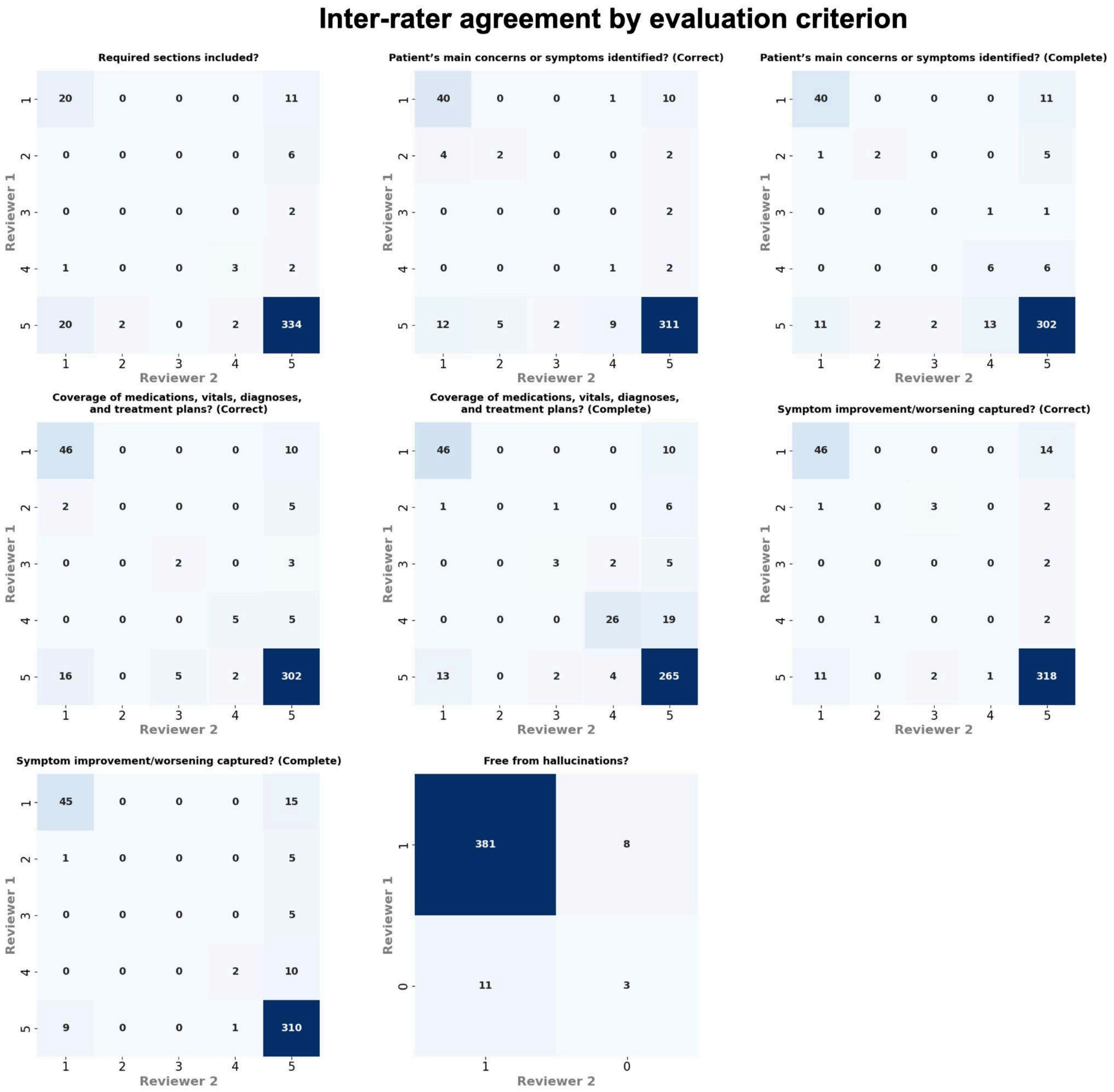


**Supplementary Figure 4: Inter-rater agreement on summary quality.** Confusion matrices compare ratings from two independent clinicians for correctness, completeness, and hallucinations. Correctness and completeness use the 1–5 rating scale; hallucinations are scored as present or absent. The concentration of high quality ratings and hallucination-free assessments should be considered when interpreting the corresponding Cohen’s κ values.

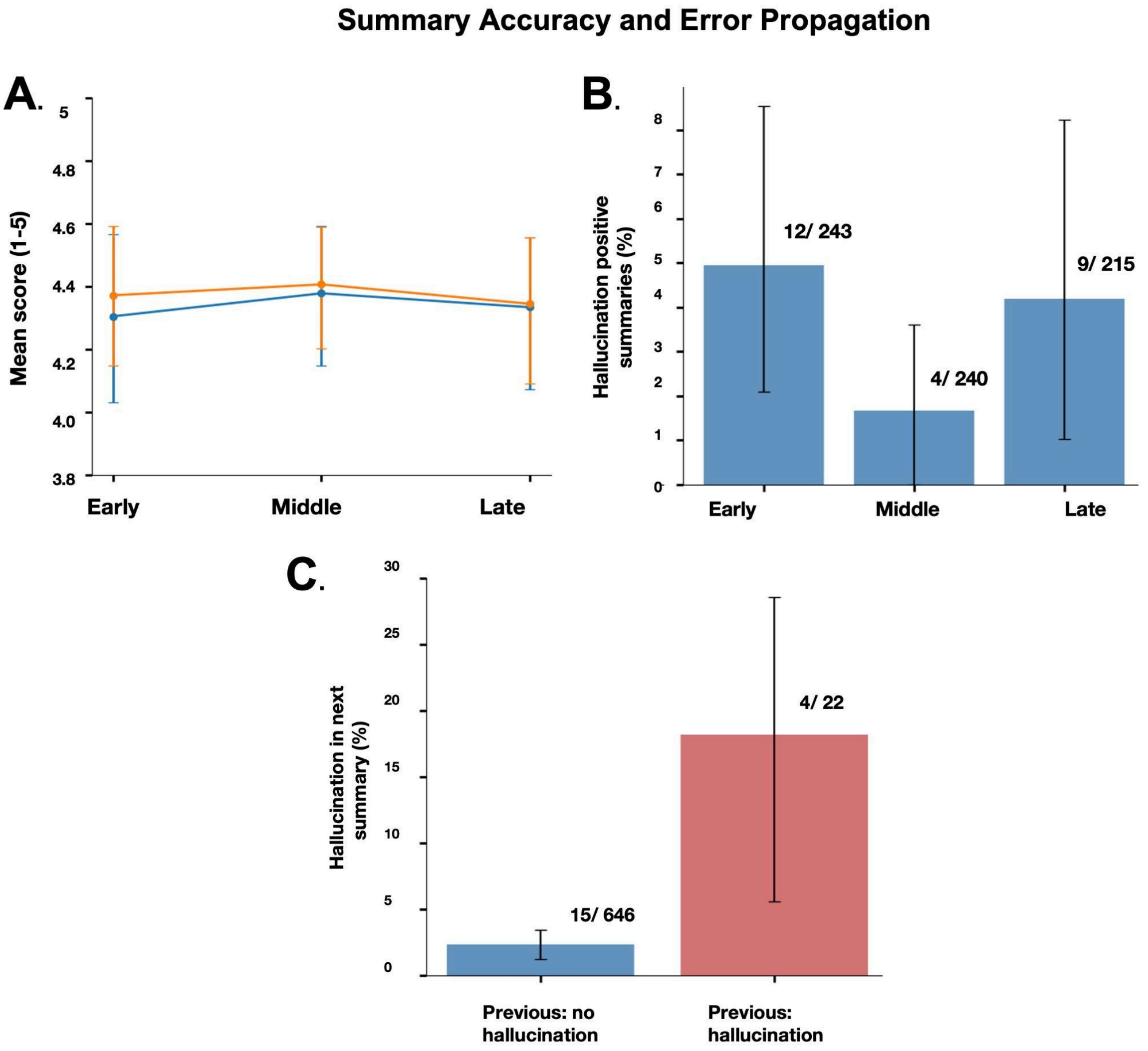


**Supplementary Figure 5: Summary accuracy and potential error propagation.** (A) Mean clinician-rated correctness (blue) and completeness (orange) across early, middle, and late summaries. (B) Percentage of hallucination-positive summaries at each stage; labels show event counts and denominators. (C) Hallucinations in the next summary occurred after 15 of 646 hallucination-free summaries (2.3%) and 4 of 22 hallucination-positive summaries (18.2%). Error bars indicate 95% confidence intervals.

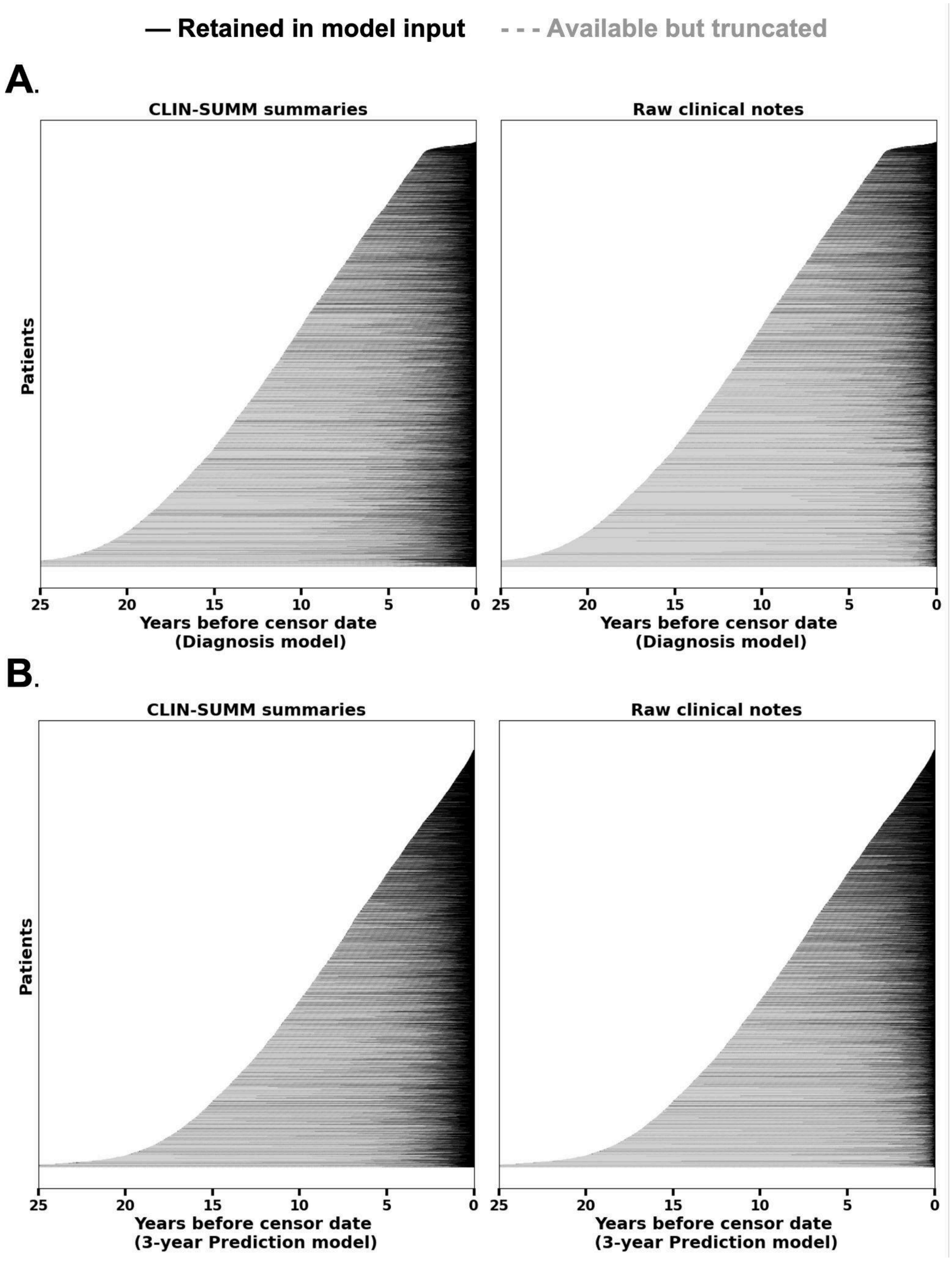


**Supplementary Figure 6: Clinical history retained at each task cutoff. (A) Diagnosis model (30-day cutoff). (B) 3-year prediction model.** Each horizontal line represents one patient, ordered by the length of available history. Black segments indicate dated CLIN-SUMM summaries (left) or raw notes (right) retained within the 8,192-token input limit; gray segments indicate available material excluded by truncation.

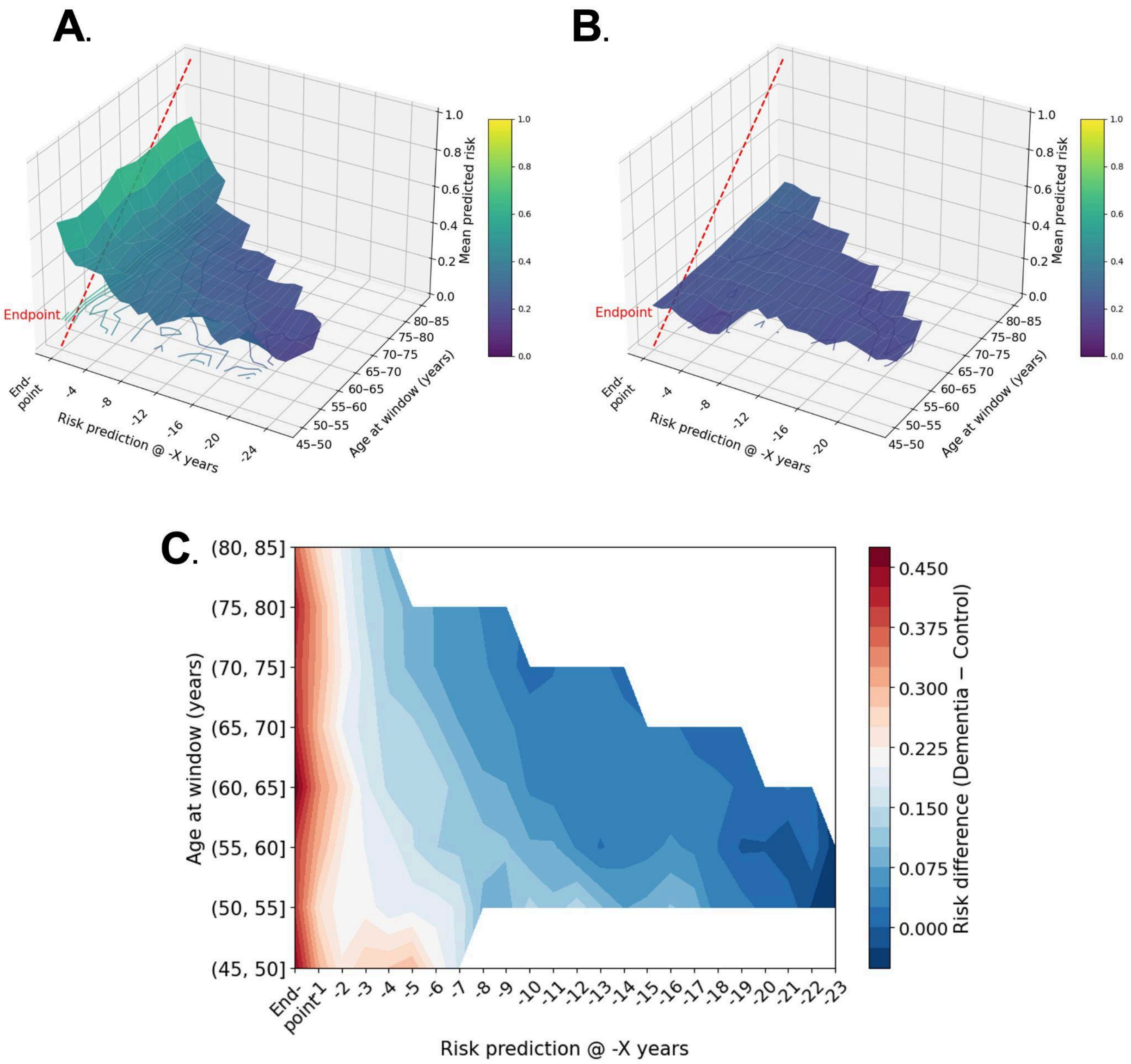


**Supplementary Figure 7:** Mean predicted dementia risk across age and time relative to the diagnostic endpoint. (A) Mean predicted dementia risk for dementia cases across age bins and time windows prior to the endpoint. (B) Mean predicted dementia risk for controls across the same age–time grid. (C) Difference surface showing the divergence in predicted risk between dementia cases and controls (dementia control). Separation between groups is greatest near the clinical endpoint across age groups.

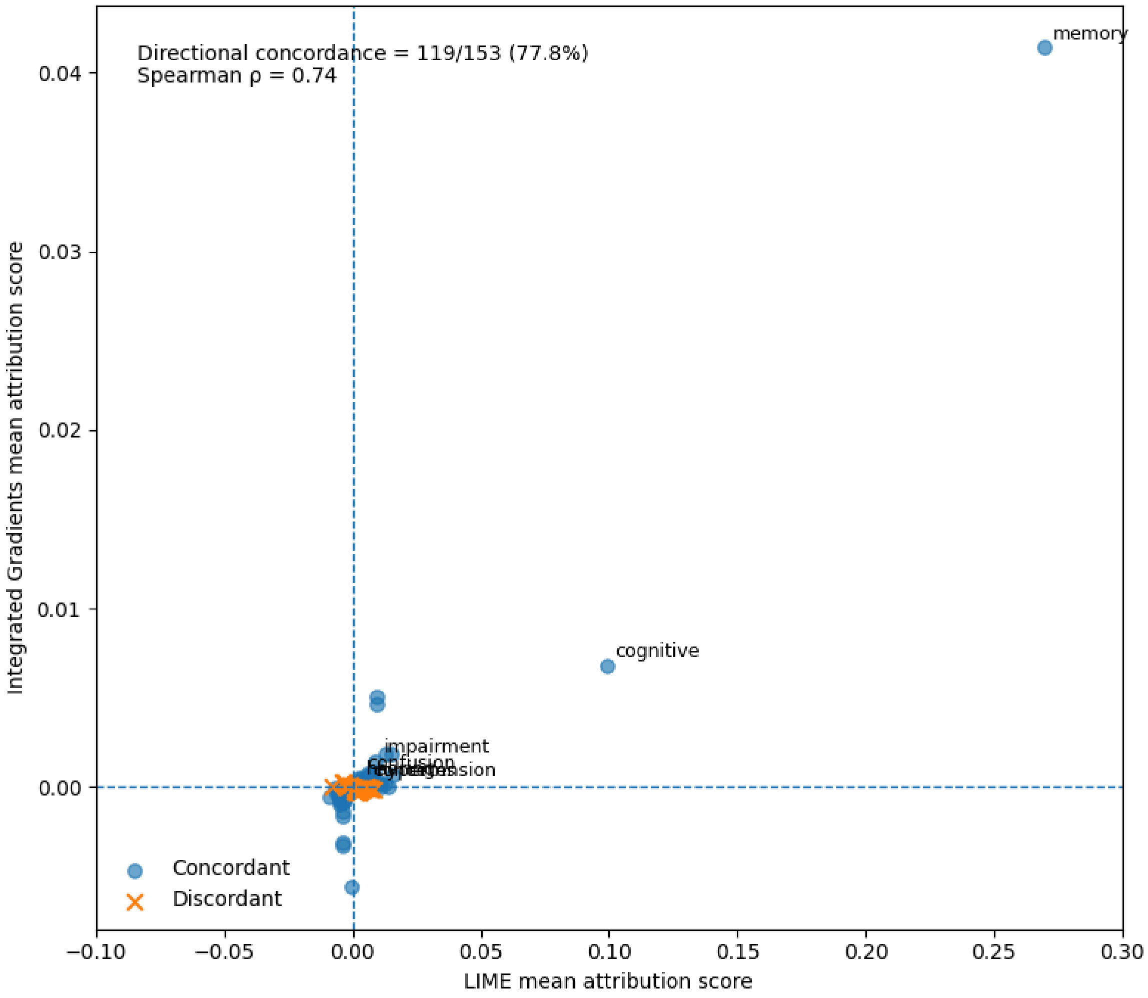


**Supplementary Figure 8: Agreement between LIME and Integrated Gradients attributions for Figure 5 features.** Each point represents a prespecified LIME feature from Figure 5, plotted against its corresponding IG attribution after aggregation from native subword tokens to words. Crosses indicate discordant attribution directions. Direction agreement was 77.8%, and Spearman rank correlation was 0.74.

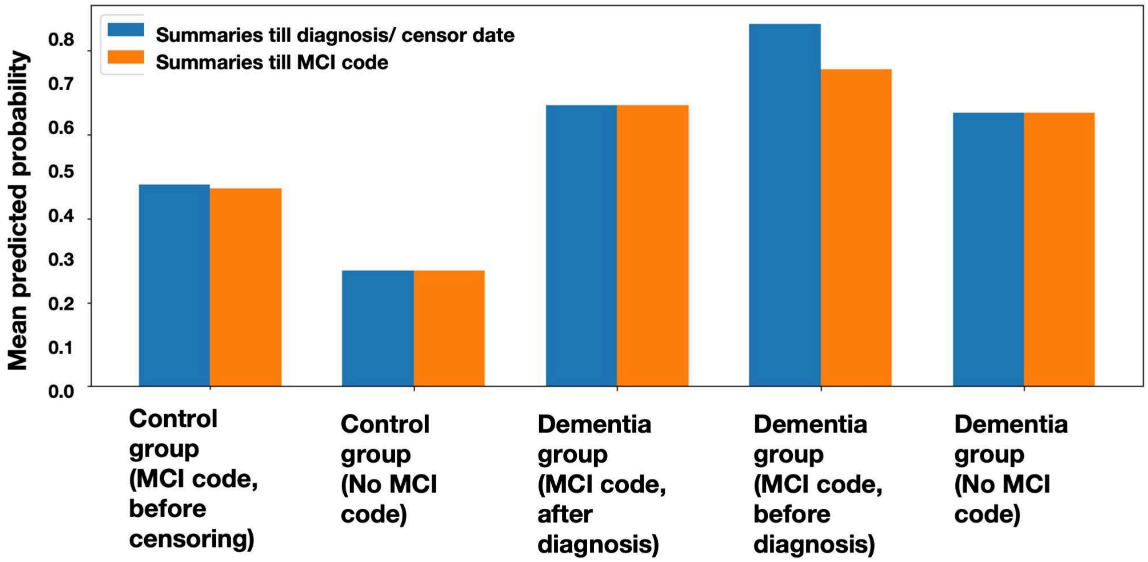


**Supplementary Figure 9:** Mean predicted dementia risk across patient subgroups under different summary truncation conditions. Subgroups include controls with a mild cognitive impairment (MCI) code before censoring, controls without an MCI code, dementia patients with an MCI code after diagnosis, dementia patients with an MCI code before diagnosis, and dementia patients without an MCI code. Blue bars show predictions using summaries up to the diagnosis or censoring date, while orange bars show predictions using summaries truncated at the first MCI code. Controls with an MCI code exhibit higher predicted risk than controls without MCI, approaching levels observed in dementia patients, while dementia groups consistently show the highest predicted probabilities.

**Supplementary Tables**

**Supplementary Table 1:** Demographic characteristics for the dementia, control and entire study cohorts. Values are reported as mean ± standard deviation for continuous variables and n (%) for categorical variables.

| Characteristic | Dementia (n=6,178) | Control (n=6,178) | All (n=12,356) |
|---|---|---|---|
| Age (years) | 75.61 ± 7.48 | 75.48 ± 7.39 | 75.54 ± 7.44 |
| Sex | | | |
| Male | 2,694 (43.61%) | 2,694 (43.61%) | 5,388 (43.61%) |
| Female | 3,484 (56.39%) | 3,484 (56.39%) | 6,968 (56.39%) |
| Race | | | |
| White | 5,149 (83.34%) | 5,206 (84.27%) | 10,355 (83.81%) |
| Black | 303 (4.90%) | 249 (4.03%) | 552 (4.47%) |
| Asian | 164 (2.65%) | 260 (4.21%) | 424 (3.43%) |
| Other | 562 (9.10%) | 463 (7.49%) | 1,025 (8.29%) |
| Average number of Notes | | | |
| Overall | 260 ± 203 | 192 ± 158 | 226 ± 185 |
| Before diagnosis/ censor date | 167 ± 162 | 144 ± 137 | 156 ± 150 |
| Average Years of Data | | | |
| Overall | 16 ± 6 | 14 ± 6 | 15 ± 6 |
| Before diagnosis/ censor date | 11 ± 5 | 11 ± 5 | 11 ± 5 |

**Supplementary Table 2:** The structured summary evaluation questionnaire used by clinicians evaluates correctness, completeness and hallucinations, comparing each individual structured summary with its corresponding raw note and all previous raw notes, implicitly including temporal coherence.

| Question | Metric | Scoring |
|---|---|---|
| Does the summary include all required sections if related information was present in the note? | Completeness | 1–5 |
| Does the summary clearly identify the patient's main concerns or symptoms? | Correctness<br>Completeness | 1–5<br>1–5 |
| Are listed medications, vitals, diagnoses, and treatment plans appropriately covered? | Correctness<br>Completeness | 1–5<br>1–5 |
| Does the summary accurately reflect any improvement, worsening, or resolution of prior complaints? | Correctness<br>Completeness | 1–5<br>1–5 |
| Is the summary free from hallucinations or invented content not supported by the source documentation? | Accuracy | Yes/No |

**Supplementary Table 3:** Validation results of CLIN-SUMM summaries for a sample of N=698 notes (corresponding to 30 patients) from the pilot set using the structured questionnaire. Inter-rater agreement scores are for a subset of N=403 notes (corresponding to 15 patients) evaluated by an independent second clinician.

| Validation Question | Score (/5 or %) | Inter-rater Agreement (%) [95% CI] | Cohen's κ |
|---|---|---|---|
| Required sections included? | | | |
| Completeness | 4.72 ± 0.97 | 88.6 [85.1 - 91.3] | 0.455 |
| Patient's main concerns or symptoms identified? | | | |
| Correctness | 4.70 ± 1.03 | 87.8 [84.3 - 90.7] | 0.594 |
| Completeness | 4.68 ± 1.02 | 86.8 [83.2 - 89.8] | 0.591 |
| Coverage of medications, vitals, diagnoses, and treatment plans? | | | |
| Correctness | 4.67 ± 1.03 | 88.1 [84.6 - 90.9] | 0.636 |
| Completeness | 4.54 ± 1.05 | 84.4 [80.5 - 87.6] | 0.642 |
| Symptom improvement/worsening captured? | | | |
| Correctness | 4.71 ± 1.01 | 90.3 [87.0 - 92.8] | 0.663 |
| Completeness | 4.65 ± 1.03 | 88.6 [85.1 - 91.3] | 0.619 |
| Free from hallucinations? | 96% | 95.3 [92.8 - 97.0] | 0.216 |

**Supplementary Table 4: Inter-rater agreement for clinical summary assessments.** Two clinicians independently rated 403 summaries from 15 patients. Ordinal ratings used a 1–5 scale. Exact agreement and unweighted Cohen's κ assess identical scores, while quadratic weighted κ accounts for the size of differences between ordinal scores. Rows labeled "≥4" dichotomize ratings as adequate (4–5) or inadequate (1–3). For hallucination assessment, positive agreement refers to ratings of *free from hallucinations* and negative agreement to ratings identifying hallucinations.

| Evaluation criterion | Scoring | Inter-rater Agreement (%) [95% CI] | Cohen's κ | Quadratic weighted κ | Positive Agreement (%) | Negative Agreement (%) |
|---|---|---|---|---|---|---|
| Required sections included? | | | | | | |
| Completeness | Original | 88.6 [85.1 - 91.3] | 0.455 | 0.470 | - | - |
| | ≥4 | 89.6 [86.2 - 92.2] | 0.430 | - | 94.199 | 48.780 |
| Patient's main concerns or symptoms identified? | | | | | | |
| Correctness | Original | 87.8 [84.3 - 90.7] | 0.594 | 0.711 | - | - |
| | ≥4 | 91.6 [88.4 - 93.9] | 0.680 | - | 95.000 | 73.016 |
| Completeness | Original | 86.8 [83.2 - 89.8] | 0.591 | 0.703 | - | - |
| | ≥4 | 91.8 [88.7 - 94.1] | 0.675 | - | 95.197 | 72.269 |
| Coverage of medications, vitals, diagnoses, and treatment plans? | | | | | | |
| Correctness | Original | 88.1 [84.6 - 90.9] | 0.636 | 0.702 | - | - |
| | ≥4 | 90.3 [87.0 - 92.8] | 0.661 | - | 94.153 | 71.942 |
| Completeness | Original | 84.4 [80.5 - 87.6] | 0.642 | 0.707 | - | - |
| | ≥4 | 90.6 [87.3 - 93.1] | 0.672 | - | 94.294 | 72.857 |
| Symptom improvement/worsening captured? | | | | | | |
| Correctness | Original | 90.3 [87.0 - 92.8] | 0.663 | 0.732 | - | - |
| | ≥4 | 92.1 [89.0 - 94.3] | 0.710 | - | 95.252 | 75.758 |
| Completeness | Original | 88.6 [85.1 - 91.3] | 0.619 | 0.715 | - | - |
| | ≥4 | 91.6 [88.4 - 93.9] | 0.681 | - | 95.000 | 73.016 |
| Free from hallucinations? | Original | 95.3 [92.8 - 97.0] | 0.216 | - | 97.567 | 24.000 |
| Overall Correct | Original | 88.8 [86.8 - 90.4] | 0.631 | 0.715 | - | - |
| | ≥4 | 91.3 [89.6 - 92.8] | 0.684 | - | 94.805 | 73.552 |
| Overall Complete | Original | 87.1 [85.4 - 88.6] | 0.598 | 0.666 | - | - |
| | ≥4 | 90.9 [89.4 - 92.2] | 0.632 | - | 94.668 | 68.522 |

**Supplementary Table 5: Summary quality across patient histories and hallucination transitions.** (A) Clinician ratings and hallucination rates for early, middle, and late summaries from 30 patients (698 summaries), plus the rate in the next summary conditional on the previous summary's hallucination status. (B) Within-patient change from early to late, calculated as late minus early. (C) Risk difference and risk ratio for a hallucination following a hallucination-positive versus hallucination-free summary.

#### A. Longitudinal summary quality and hallucination rates

| Analysis | Group | N | Hallucinations | Correctness | Completeness | Hallucination (%) |
|---|---|---|---|---|---|---|
| Accuracy across stages | Early | 243 | 12 | 4.30 [4.03, 4.57] | 4.37 [4.15, 4.59] | 4.9 [2.1, 8.5] |
| Accuracy across stages | Middle | 240 | 4 | 4.38 [4.15, 4.59] | 4.41 [4.20, 4.59] | 1.7 [0.0, 3.6] |
| Accuracy across stages | Late | 215 | 9 | 4.33 [4.07, 4.55] | 4.34 [4.09, 4.56] | 4.2 [1.0, 8.2] |
| One-step transition | Previous: hallucination-free | 646 | 15 | - | - | 2.3 [1.2, 3.5] |
| One-step transition | Previous: hallucination-positive | 22 | 4 | - | - | 18.2 [5.6, 28.6] |

#### B. Paired patient–level Early vs Late comparison

| Metric | Patients | Early mean | Late mean | Difference | p-value |
|---|---|---|---|---|---|
| Correctness | 30 | 4.31 | 4.33 | 0.02 | 0.59 |
| Completeness | 30 | 4.38 | 4.34 | -0.04 | 0.96 |
| Hallucinations rate | 30 | 0.05 | 0.05 | 0.00007 | 0.86 |

#### C. Transition effect estimates

| Statistic | Estimate | 95% CI lower | 95% CI upper |
|---|---|---|---|
| Risk difference (percentage points) | 15.86 | 3.49 | 26.30 |
| Risk ratio | 7.83 | 2.43 | 17.32 |

**Supplementary Table 6. Retained history for the Diagnosis model (30-day cutoff).** Truncation frequency, available and retained history, and token retention by case status and eligible note count. Years are medians [interquartile ranges]; older-material retention is reported among patients with such material available.

| Group | Eligible raw notes | Patients, n | Available history, years | Summary truncated, % | Raw notes truncated, % | Summary: oldest retained, years | Raw: oldest retained, years | Summary: >5y material retained | Raw: >5y material retained | Summary tokens retained, median % | Raw tokens retained, median % |
|---|---|---|---|---|---|---|---|---|---|---|---|
| Case | All | 6178 | 11.7 [7.4–16.2] | 89.1 | 90.1 | 1.9 [1.0–3.6] | 0.9 [0.4–2.3] | 939/5470 (17.2%) | 612/5463 (11.2%) | 22.3 | 15.6 |
| Control | All | 6178 | 11.1 [7.2–15.5] | 86.2 | 88.6 | 2.3 [1.1–4.5] | 1.0 [0.4–3.0] | 1278/5422 (23.6%) | 772/5417 (14.3%) | 25.4 | 16.9 |
| Case | 1–50 | 1270 | 6.3 [4.2–10.0] | 47.1 | 52.2 | 4.7 [3.2–7.4] | 3.9 [2.1–6.4] | 583/817 (71.4%) | 465/813 (57.2%) | 100 | 95.7 |
| Case | 51–100 | 1333 | 9.8 [6.5–13.8] | 99.8 | 99.5 | 2.4 [1.6–4.0] | 1.7 [0.7–2.9] | 222/1174 (18.9%) | 116/1172 (9.9%) | 37 | 31.8 |
| Case | 101–200 | 1799 | 12.3 [9.1–16.4] | 100 | 100 | 1.6 [1.0–2.7] | 0.8 [0.4–1.5] | 110/1721 (6.4%) | 23/1720 (1.3%) | 19.1 | 13.9 |
| Case | >200 | 1776 | 15.6 [12.3–19.0] | 100 | 100 | 1.0 [0.6–1.7] | 0.3 [0.2–0.6] | 24/1758 (1.4%) | 8/1758 (0.5%) | 8.8 | 4.4 |
| Control | 1–50 | 1588 | 6.7 [4.6–10.0] | 46.6 | 55.9 | 5.3 [3.6–8.2] | 4.2 [2.2–6.8] | 855/1106 (77.3%) | 630/1102 (57.2%) | 100 | 86.8 |
| Control | 51–100 | 1402 | 9.6 [6.5–13.7] | 99.9 | 99.7 | 2.7 [1.6–4.5] | 1.6 [0.7–3.0] | 271/1211 (22.4%) | 120/1210 (9.9%) | 37.3 | 29.2 |
| Control | 101–200 | 1723 | 12.5 [9.0–16.9] | 100 | 100 | 1.7 [0.9–3.0] | 0.7 [0.4–1.4] | 129/1648 (7.8%) | 19/1648 (1.2%) | 18.6 | 12.3 |
| Control | >200 | 1465 | 15.0 [11.9–18.7] | 100 | 100 | 1.0 [0.5–1.7] | 0.3 [0.2–0.6] | 23/1457 (1.6%) | 3/1457 (0.2%) | 8.9 | 4.5 |

**Supplementary Table 7. Retained history for the 3-year Prediction model.** Truncation frequency, available and retained history, and token retention by case status and eligible note count. Years are medians [interquartile ranges]; older-material retention is reported among patients with such material available.

| Group | Eligible raw notes | Patients, n | Available history, years | Summary truncated, % | Raw notes truncated, % | Summary: oldest retained, years | Raw: oldest retained, years | Summary: >5y material retained | Raw: >5y material retained | Summary tokens retained, median % | Raw tokens retained, median % |
|---|---|---|---|---|---|---|---|---|---|---|---|
| Case | All | 6140 | 11.8 [7.5–16.3] | 73 | 74 | 5.0 [4.0–6.7] | 4.4 [3.5–6.1] | 3021/5466 (55.3%) | 2328/5463 (42.6%) | 39.5 | 35.1 |
| Control | All | 6178 | 11.1 [7.2–15.5] | 68.7 | 71.2 | 5.2 [4.1–7.0] | 4.5 [3.6–6.2] | 3315/5421 (61.2%) | 2471/5417 (45.6%) | 46.7 | 39.5 |
| Case | 1–50 | 2376 | 7.0 [4.9–10.4] | 30.2 | 33.2 | 6.1 [4.6–8.7] | 5.8 [4.3–8.3] | 1596/1729 (92.3%) | 1477/1727 (85.5%) | 100 | 100 |
| Case | 51–100 | 1376 | 11.7 [9.1–14.9] | 99.9 | 99.3 | 5.4 [4.5–6.7] | 4.8 [4.1–6.2] | 805/1354 (59.5%) | 634/1353 (46.9%) | 39.6 | 37.1 |
| Case | 101–200 | 1336 | 14.8 [11.8–18.1] | 100 | 100 | 4.6 [4.0–5.4] | 3.9 [3.5–4.5] | 477/1331 (35.8%) | 189/1331 (14.2%) | 19.9 | 15.9 |
| Case | >200 | 1052 | 17.2 [14.4–19.9] | 100 | 100 | 3.9 [3.6–4.5] | 3.4 [3.2–3.6] | 143/1052 (13.6%) | 28/1052 (2.7%) | 9.6 | 5.9 |
| Control | 1–50 | 2733 | 7.2 [4.9–10.2] | 29.2 | 35.1 | 6.3 [4.6–8.8] | 5.8 [4.3–8.3] | 1870/2004 (93.3%) | 1694/2000 (84.7%) | 100 | 100 |
| Control | 51–100 | 1331 | 11.6 [9.0–15.1] | 99.9 | 99.2 | 5.5 [4.5–6.9] | 4.8 [3.9–6.0] | 811/1308 (62.0%) | 596/1308 (45.6%) | 38.9 | 35.3 |
| Control | 101–200 | 1311 | 14.8 [11.9–18.2] | 100 | 100 | 4.7 [4.0–5.6] | 3.8 [3.4–4.4] | 509/1306 (39.0%) | 166/1306 (12.7%) | 19.7 | 15.1 |
| Control | >200 | 803 | 16.9 [13.9–20.0] | 100 | 100 | 3.9 [3.6–4.6] | 3.4 [3.2–3.6] | 125/803 (15.6%) | 15/803 (1.9%) | 9.8 | 6.1 |

**Supplementary Table 8: Performance of summary and raw-note representations.** TF-IDF and Clinical ModernBERT were evaluated for Diagnosis (30 days before the first recorded dementia diagnosis) and 3-year prediction tasks. Clinical ModernBERT inputs were limited to 8,192 tokens, retaining the most recent text when necessary; TF-IDF used the same selected text.

| Task and input | Model | *AUROC* | *AUPRC* | *F1 score* | *Precision* | *Recall* |
|---|---|---|---|---|---|---|
| **Diagnosis (Summaries as input)** | **TF-IDF** | 0.86 [0.85 - 0.86] | 0.86 [0.85 - 0.87] | 0.77 | 0.81 | 0.73 |
| | **Clinical ModernBERT** | 0.84 [0.83 - 0.85] | 0.86 [0.85 - 0.86] | 0.75 | 0.82 | 0.69 |
| **Diagnosis (Raw notes as input)** | **TF-IDF** | 0.84 [0.83 - 0.85] | 0.84 [0.83 - 0.85] | 0.76 | 0.77 | 0.74 |
| | **Clinical ModernBERT** | 0.83 [0.82 - 0.84] | 0.84 [0.83 - 0.85] | 0.75 | 0.78 | 0.73 |
| **3-year Prediction (Summaries as input)** | **TF-IDF** | 0.65 [0.64 - 0.66] | 0.66 [0.64 - 0.67] | 0.59 | 0.62 | 0.57 |
| | **Clinical ModernBERT** | 0.66 [0.65 - 0.67] | 0.67 [0.66 - 0.69] | 0.58 | 0.64 | 0.54 |
| **3-year Prediction (Raw notes as input)** | **TF-IDF** | 0.65 [0.64 - 0.66] | 0.64 [0.63 - 0.66] | 0.58 | 0.61 | 0.56 |
| | **Clinical ModernBERT** | 0.63 [0.62 - 0.64] | 0.64 [0.62 - 0.65] | 0.55 | 0.61 | 0.50 |

**Supplementary Table 9:** Performance of the Clinical-ModernBERT models in the test cohort using CLIN-SUMM summaries for dementia diagnosis and 3-year prediction, compared to baseline logistic regression models that input age–sex, age–sex–comorbidities, age–sex–comorbidities–vitals, or age–sex–comorbidities–vitals-utilization stats. The best results for each task are bolded. AUROC and AUPRC show the result with [95% CI].

| **Diagnosis** | | | | | | |
|---|---|---|---|---|---|---|
| **All test subjects (N=8942)** | | | | | | |
| **Metric** | **Age–Sex** | **Age–Sex–Comorbidities** | **Clinical ModernBERT on Summaries** | **Age–Sex–Utilization stats** | **Age–Sex–Comorbidities–Utilization stats** | **Age–Sex–Comorbidities–Utilization stats–Clinical ModernBERT on Summaries scores** |
| Model | Logistic | Logistic | Transformer | Logistic | Logistic | Logistic |
| AUROC | 0.51[0.5–0.52] | 0.63 [0.62–0.64] | **0.84 [0.83–0.85]** | 0.55 [0.53–0.56] | 0.63 [0.62–0.64] | **0.84 [0.83–0.84]** |
| AUPRC | 0.51 [0.5–0.53] | 0.63 [0.62–0.65] | **0.86 [0.85–0.86]** | 0.55 [0.54 - 0.57] | 0.64 [0.62 - 0.65] | **0.84 [0.83–0.85]** |
| F1 Score | 0.54 | 0.56 | **0.75** | 0.46 | 0.56 | **0.76** |
| Precision | 0.5 | 0.62 | **0.82** | 0.55 | 0.62 | **0.81** |
| Recall | 0.6 | 0.51 | **0.69** | 0.39 | 0.51 | **0.72** |
| **Test subjects with vitals (N=5789)** | | | | | | |
| **Metric** | **Age–Sex** | **Age–Sex–Comorbidities–Vitals** | **Clinical ModernBERT on Summaries** | **Age–Sex–Utilization stats** | **Age–Sex–Comorbidities–Vitals–Utilization stats** | **Age–Sex–Comorbidities–Vitals–Utilization stats–Clinical ModernBERT on Summaries scores** |
| Model | Logistic | Logistic | Transformer | Logistic | Logistic | Logistic |
| AUROC | 0.5 [0.49–0.52] | 0.65 [0.63–0.66] | **0.86 [0.85–0.87]** | 0.56 [0.54 –0.57] | 0.65 [0.63 - 0.66] | **0.86 [0.85–0.87]** |
| AUPRC | 0.5 [0.49–0.53] | 0.65 [0.63–0.67] | **0.87 [0.86–0.88]** | 0.56 [0.54–0.58] | 0.65 [0.63–0.67] | **0.86 [0.84–0.87]** |
| F1 Score | 0.53 | 0.58 | **0.77** | 0.49 | 0.58 | **0.79** |
| Precision | 0.5 | 0.62 | **0.83** | 0.56 | 0.62 | **0.82** |
| Recall | 0.57 | 0.54 | **0.73** | 0.43 | 0.55 | **0.76** |
| **3-year Prediction** | | | | | | |
| **All test subjects (N=8906)** | | | | | | |
| **Metric** | **Age–Sex** | **Age–Sex–Comorbidities** | **Clinical ModernBERT on Summaries** | **Age–Sex–Utilization stats** | **Age–Sex–Comorbidities–Utilization stats** | **Age–Sex–Comorbidities–Utilization stats–Clinical ModernBERT on Summaries scores** |
| Model | Logistic | Logistic | Transformer | Logistic | Logistic | Logistic |
| AUROC | 0.51 [0.5–0.52] | 0.59 [0.57–0.6] | **0.66 [0.65–0.67]** | 0.54 [0.53–0.55] | 0.59[0.58–0.6] | **0.66 [0.65–0.67]** |
| AUPRC | 0.51 [0.5–0.53] | 0.59 [0.58–0.61] | **0.67 [0.66–0.69]** | 0.54 [0.53–0.56] | 0.59[0.58–0.61] | **0.67 [0.66–0.69]** |
| F1 Score | 0.53 | 0.51 | **0.58** | 0.43 | 0.51 | **0.59** |
| Precision | 0.5 | 0.58 | **0.64** | 0.54 | 0.58 | **0.64** |
| Recall | 0.57 | 0.45 | **0.54** | 0.36 | 0.46 | **0.54** |
| **Test subjects with vitals (N=4761)** | | | | | | |

| **Metric** | **Age–Sex** | **Age–Sex–Comorbidities–Vitals** | **Clinical ModernBERT on Summaries** | **Age–Sex–Utilization stats** | **Age–Sex–Comorbidities–Vitals–Utilization stats** | **Age–Sex–Comorbidities–Vitals–Utilization stats–Clinical ModernBERT on Summaries scores** |
|---|---|---|---|---|---|---|
| Model | Logistic | Logistic | Transformer | Logistic | Logistic | Logistic |
| AUROC | 0.5 [0.49–0.52] | 0.6 [0.58–0.61] | **0.68 [0.67–0.7]** | 0.54 [0.53–0.56] | 0.6 [0.58–0.61] | **0.69 [0.67–0.7]** |
| AUPRC | 0.5 [0.48–0.52] | 0.61 [0.58–0.63] | **0.69 [0.67–0.71]** | 0.55[0.53–0.57] | 0.6 [0.58–0.62] | **0.68 [0.66–0.7]** |
| F1 Score | 0.52 | 0.54 | **0.61** | 0.47 | 0.54 | **0.61** |
| Precision | 0.49 | 0.57 | **0.65** | 0.54 | 0.57 | **0.65** |
| Recall | 0.54 | 0.51 | **0.57** | 0.42 | 0.51 | **0.58** |

**Supplementary Table 10. Documentation volume and history availability by case status and task.** Eligible note counts, available summary tokens, observation time, and summary-input truncation rates in the full cohort at the diagnosis and 3-year prediction cutoffs. Values are median [25th–75th percentile] or percentages. Truncation indicates inputs exceeding 8,192 tokens.

| Task | Group | Patients | Eligible notes, median [IQR] | Available summary tokens, median [IQR] | Observation time, years, median [IQR] | Summary input truncated |
|---|---|---|---|---|---|---|
| Diagnosis | Cases | 6,178 | 122 [60–221] | 36661 [17626–67800] | 11.7 [7.4–16.2] | 89.10% |
| | Controls | 6,178 | 105 [49–195] | 32253 [14305–60343] | 11.1 [7.2–15.5] | 86.20% |
| 3-year Prediction | Cases | 6,140 | 72 [26–153] | 20701 [7193–45374] | 11.8 [7.5–16.3] | 73.00% |
| | Controls | 6,178 | 60 [21–133] | 17544 [5739–40136] | 11.1 [7.2–15.5] | 68.70% |

**Supplementary Table 11. Performance of utilization-based models and Clinical ModernBERT on CLIN-SUMM summaries**. AUROC and AUPRC are reported with 95% confidence intervals for the held-out diagnosis (30-day; N=8,942) and 3-year prediction (N=8,906) test sets. Utilization variables comprise eligible note count, available history in years, and available summary token count before each cutoff. Test sets have approximately equal numbers of cases and controls; AUPRC therefore reflects this matched evaluation setting.

| | Model | AUROC | AUPRC |
|---|---|---|---|
| Diagnosis, N=8,942 | Utilization only: note count, available years, available tokens | 0.55 [0.54 - 0.56] | 0.55 [0.54 - 0.56] |
| | Age–sex + utilization stats | 0.55 [0.53–0.56] | 0.55 [0.54–0.57] |
| | Age–sex + comorbidities + utilization stats | 0.63 [0.62–0.64] | 0.64 [0.62–0.65] |
| | Clinical ModernBERT on summaries | 0.84 [0.83–0.85] | 0.86 [0.85–0.86] |
| 3-year Prediction, N=8,906 | Utilization only: note count, available years, available tokens | 0.54 [0.52 - 0.55] | 0.54 [0.53 - 0.56] |
| | Age–sex + utilization stats | 0.54 [0.53–0.55] | 0.54 [0.53–0.56] |
| | Age–sex + comorbidities + utilization stats | 0.59 [0.58–0.60] | 0.59 [0.58–0.61] |
| | Clinical ModernBERT on summaries | 0.66 [0.65–0.67] | 0.67 [0.66–0.69] |

**Supplemental Table 12: Sensitivity of Clinical ModernBERT performance to the cutoff before the first recorded dementia diagnosis.** Models were trained and evaluated separately using CLIN-SUMM summaries derived from notes recorded at least 30 days, 90 days, or six months before diagnosis. Brackets indicate 95% confidence intervals.

| Metric | < 30 days before diagnosis | < 90 days before diagnosis | < 6months before diagnosis |
|---|---|---|---|
| ***AUROC*** | 0.84 [0.83 - 0.85] | 0.82 [0.82 - 0.83] | 0.78 [0.77 - 0.79] |
| ***AUPRC*** | 0.86 [0.85 - 0.86] | 0.84 [0.83 - 0.85] | 0.80 [0.78 - 0.81] |
| ***F1 score*** | 0.75 | 0.75 | 0.65 |
| ***Precision*** | 0.82 | 0.78 | 0.82 |
| ***Recall*** | 0.69 | 0.72 | 0.53 |

**Supplementary Table 13:** Time-horizon evaluation using the Diagnosis model applied to earlier cutoffs. AUROC and AUPRC show the result with [95% CI].

| Metric | 5 years | 3 years | 1 year | 6 months | 90 days | 30 days |
|---|---|---|---|---|---|---|
| ***AUROC*** | 0.62 [0.61 - 0.63] | 0.67 [0.65 - 0.68] | 0.74 [0.73 - 0.75] | 0.78 [0.77 - 0.79] | 0.81 [0.8 - 0.82] | 0.84 [0.83 - 0.85] |
| ***AUPRC*** | 0.63 [0.62 - 0.65] | 0.68 [0.66 - 0.69] | 0.76 [0.74 - 0.77] | 0.8 [0.79 - 0.81] | 0.83 [0.82 - 0.84] | 0.86 [0.85 - 0.86] |
| ***F1 Score*** | 0.35 | 0.44 | 0.6 | 0.67 | 0.71 | 0.75 |
| ***Precision*** | 0.72 | 0.73 | 0.78 | 0.8 | 0.81 | 0.82 |
| ***Recall*** | 0.23 | 0.32 | 0.49 | 0.57 | 0.64 | 0.69 |

**Supplementary Table 14:** BERTScore comparison between Qwen3- and GPT-4o-generated CLIN-SUMM summaries in the pilot cohort.

| Metric | Min | Max | Mean | Std. Dev. |
|---|---|---|---|---|
| F1 | 0.83 | 0.95 | 0.90 | 0.017 |
| Precision | 0.82 | 0.95 | 0.90 | 0.018 |
| Recall | 0.83 | 0.95 | 0.91 | 0.017 |

**Supplementary Table 15:** ICD-9 and ICD-10 diagnostic codes used to ascertain dementia.

| ICD-9 Name | Code | ICD-10 Name | Code |
|---|---|---|---|
| Senile dementia, uncomplicated | 290.0 | Vascular dementia | F01.x |
| Senile dementia with delirium | 290.10 | Vascular dementia with delusions | F01.5 |
| Senile dementia with delusional features | 290.11 | Vascular dementia with delusions (unspecified) | F01.50 |
| Senile dementia with depressive features | 290.12 | Vascular dementia with behavioral disturbance | F01.51 |
| Senile dementia with behavioral disturbance | 290.13 | Dementia in other diseases classified elsewhere | F02.x |
| Arteriosclerotic dementia | 290.20 | Dementia in other diseases classified elsewhere | F02.8 |
| Vascular dementia | 290.40 | Dementia in other diseases classified elsewhere without behavioral disturbance | F02.80 |
| Vascular dementia with behavioral disturbance | 290.21 | Dementia in other diseases with behavioral disturbance | F02.81 |
| Vascular dementia with behavioral disturbance | 290.3 | Unspecified dementia | F03.x |
| Vascular dementia with delirium | 290.41 | Unspecified dementia | F03.9 |
| Vascular dementia with delusions | 290.42 | Unspecified dementia without behavioral disturbance | F03.90 |
| Vascular dementia with depressed mood | 290.43 | Unspecified dementia with behavioral disturbance | F03.91 |
| Other specified senile psychotic conditions | 290.8 | Progressive supranuclear ophthalmoplegia | G23.1 |
| Unspecified senile psychotic condition | 290.9 | Alzheimer’s disease with early onset | G30.0 |
| Dementia in conditions classified elsewhere | 294.10 | Alzheimer’s disease with late onset | G30.1 |
| Persistent mental disorders due to conditions classified elsewhere | 294.1 | Other Alzheimer’s disease | G30.8 |
| Dementia in conditions classified elsewhere with behavioral disturbance | 294.11 | Alzheimer’s disease, unspecified | G30.9 |
| Amnestic disorder due to physiological condition | 294.20 | Frontotemporal dementia | G31.0 |
| Amnestic disorder with behavioral disturbance | 294.21 | Primary progressive aphasia | G31.01 |
| Alzheimer’s disease | 331.0 | Other frontotemporal dementia | G31.09 |

| | | | |
|---|---|---|---|
| Frontotemporal dementia (Pick's disease) | 331.1 | Senile degeneration of brain | G31.1 |
| Presenile dementia with delirium | 331.11 | Corticobasal degeneration | G31.83 |
| Presenile dementia with delusional features | 331.19 | | |
| Senile degeneration of brain | 331.2 | | |
| Dementia with Lewy bodies | 331.82 | | |

**Supplementary Table 16:** SNOMED diagnostic codes used to ascertain dementia.

| Name | SNOMED Concept IDs | SNOMED Concept codes | Name | SNOMED Concept IDs | SNOMED Concept codes |
|---|---|---|---|---|---|
| Dementia | 4182210 | 52448006 | Senile dementia with depression | 379784 | 191459006 |
| Dementia associated with another disease | 374888 | 191519005 | Senile dementia with depressive or paranoid features | 4101137 | 191457008 |
| Traumatic encephalopathy | 4047745 | 230282000 | Primary degenerative dementia of Alzheimer type, senile onset, uncomplicated | 4278830 | 66108005 |
| Cerebral degeneration presenting primarily with dementia | 4092747 | 279982005 | Multi-infarct dementia with delirium | 444091 | 10349009 |
| Alzheimer’s disease | 378419 | 26929004 | Dementia of the Alzheimer type with behavioral disturbance | 43530664 | 1581000119101 |
| Vascular dementia | 443605 | 429998004 | Mixed dementia | 43021816 | 79341000119107 |
| Dementia with behavioral disturbance | 43530666 | 1591000119103 | Multi-infarct dementia with depression | 443864 | 14070001 |
| Vascular dementia without behavioral disturbance | 37109056 | 16276361000119109 | Senile dementia with delirium | 376946 | 191461002 |
| Senile dementia | 4048875 | 15662003 | Presenile dementia with delusions | 44782771 | 31081000119101 |
| Presenile dementia | 378125 | 12348006 | Senile dementia of the Lewy body type | 4196433 | 312991009 |
| Primary degenerative dementia of Alzheimer type, senile onset | 4220313 | 416975007 | Senile dementia with delusion | 380986 | 371024007 |
| Uncomplicated senile dementia | 375791 | 191449005 | Mild dementia | 762497 | 428051000124108 |
| Frontotemporal dementia | 4043378 | 230270009 | Multi-infarct dementia with delusions | 443790 | 25772007 |
| Primary degenerative dementia of Alzheimer type, presenile onset | 4218017 | 416780008 | Lewy body dementia with behavioral disturbance | 44782763 | 135811000119107 |
| Vascular dementia with behavioral disturbance | 37018688 | 288631000119104 | Presenile dementia with delirium | 381832 | 191452002 |
| Diffuse Lewy body disease | 380701 | 80098002 | Primary degenerative dementia of Alzheimer type, presenile onset, uncomplicated | 4277444 | 6475002 |
| Multi-infarct dementia | 379778 | 56267009 | Dementia of frontal lobe type | 441002 | 278857002 |
| Uncomplicated presenile dementia | 376085 | 191451009 | Mixed cortical and subcortical vascular dementia | 4046090 | 230287006 |
| Multi-infarct dementia, uncomplicated | 377254 | 70936005 | Psychoactive substance-induced organic dementia | 4009647 | 111480006 |
| Drug-induced dementia | 376095 | 191493005 | Subcortical dementia | 42538857 | 762707000 |
| Dementia associated with alcoholism | 378726 | 281004 | Subcortical vascular dementia | 4047747 | 230286002 |
| Presenile dementia with depression | 377527 | 191455000 | Dementia due to disorder of central nervous system | 37110513 | 724776007 |

| | | | | | |
|---|---|---|---|---|---|
| Behavioral and psychological symptoms of dementia | 35608576 | 10171000132106 | Dementia due to Huntington chorea | 40483103 | 442344002 |
| Inhalant-induced persisting dementia | 4139421 | 32875003 | | | |

**Supplementary Table 17:** Performance metrics by distance bin, including similarity, recall, and information loss, used to determine similarity threshold for Jaccard distance (In the pilot set of 1500).

| Distance Bin | Mean Similarity (%) | Mean Entity Recall (%) | Info Lost (%) | # Pairs |
|---|---|---|---|---|
| (0.00 – 0.01] | 99.97 | 99.18 | 0.82 | 1006 |
| (0.01 – 0.02] | 98.53 | 96.68 | 3.32 | 111 |
| (0.02 – 0.03] | 97.45 | 96.17 | 3.83 | 148 |
| (0.03 – 0.04] | 96.46 | 94.12 | 5.88 | 182 |
| (0.04 – 0.05] | 95.50 | 94.31 | 5.69 | 211 |
| (0.05 – 0.06] | 94.53 | 92.77 | 7.23 | 239 |
| (0.06 – 0.07] | 93.50 | 93.02 | 6.98 | 237 |
| (0.07 – 0.08] | 92.50 | 91.81 | 8.19 | 316 |
| (0.08 – 0.09] | 91.50 | 91.13 | 8.87 | 253 |
| (0.09 – 0.10] | 90.52 | 90.58 | 9.42 | 278 |

**Supplementary Table 18:** Diagnostic codes used to extract comorbidities from the EHR.

| Comorbidity | ICD Codes | SNOMED Concept IDs | SNOMED Concept codes |
|---|---|---|---|
| Alcohol use disorder | F10.10, F10.11, F10.14, F10.19, F10.12*, F10.13*, F10.15*, F10.18*, 305.0, 305.00–305.03 | 433753 | 15167005 |
| Anxiety disorders | F41.1, F41.3, F41.8, F41.9, 300.0, 300.00, 300.02, 300.09 | 36684319, 442077, 4113821, 434613 | 782501005, 197480006, 198288003, 21897009 |
| Delirium | 293.0, 293.1, F05 | 373995, 379779 | 2776000, 191507002 |
| Hyperlipidemia | E78.2, E78.4, E78.5, 272.2, 272.4 | 432867, 438720 | 55822004, 267434003 |
| Dizziness | R42, 780.4 | 433316 | 271789005 |
| Dyspnea | R06.0, R06.00, R06.09 | 312437 | 267036007 |
| Osteoporosis | M81.0, M81.6, M81.8, 733.0, 733.00–733.03, 733.09 | 80824, 80502, 4004623, 81390, 77365 | 18040001, 64859006, 203435007, 3345002, 53174001 |
| Syncope | R55, 780.2, 992.1, G90.01, T67.1XXA, T67.1XXD, T67.1XXS | 436392, 140586, 4240219, 4206148 | 51723007, 89797005, 58077008, 309585006 |
| Parkinson’s disease | G20, G21.2, G21.3, G21.4, G21.8, G21.9, 332, 332.1 | 381270, 4140090, 4064308, 374013, 4046093 | 49049000, 32798002, 19972008, 230292008, 230296006 |
| Weight loss / anorexia | R63.0, R63.4, F50.0–F50.02, 783.0, 307.1 | 435928, 436675, 4269485, 4300305, 4229881 | 267024001, 56882008, 63393005, , 77675002, 89362005 |
| Visual impairment | H53.121–H53.133, H53.139, H54.3, H54.60–H54.62, H54.7, 368.11–368.12, 369.3, 369.8, 369.9 | 45773067, 45757571, 4288371, 4288370, 377556, 372627, 4265433 | 8771000119104, 218951000119100, 397542006, 397541004, 15203004, 36994005, 397540003 |
| Hearing impairment | H90.*, H91.*, 389.*, 388.12, 388.2 | 374053, 374366, 374367, 375826, 376131, 376417, 377888, 377889, 378442, 379832, 380734, 380742, 433495, 436421, 440422, 443577, 443606, 443608, 444291, 4110815, 43021778, 43021779 | 79471008, 60700002, 95820000, 68467004, 15190000, 61743004, 44057004, 15188001, 2061000, 77507001, 47111006, 3820005, 194417009, 194429000, 73415002, 428887009, 430977001, 430985005, 85571008, 194424005, 473423001, 473424007 |
| Hypertension | 401.*, 405.*, 997.91, 572.3, 459.3*, 416.0, 365.04, 348.2, I10, I15.*, I27.*, I87.*, I97.3, K76.6, H40.05*, G93.2 | 192679,192680, 312648, 312902, 314958, 317895, 317898, 318437, 319826, 320128, 381290, 443771, 4013643, 4110947, 4110948, 4249016, 4311246, 4313767, 4339214, 44782429, 44782690, 44782715 | 75150001, 34742003, 1201005, 68267002, 194785008, 123799005, 78975002, 89242004, 31992008, 59621000, 4210003, 28119000, 11399002, 194783001, 194788005, 73410007, 86041002, 423674003, 88223008, 104931000119100, 129161000119100, 153811000119105 |

**Supplementary Table 19:** SNOMED codes (measurement table) used to extract vital signs and laboratory values.

| Measurement | Snomed Concept IDs | Snomed Concept Codes |
|---|---|---|
| Systolic blood pressure | 3004249 | 8480-6 |
| Diastolic blood pressure | 3012888 | 8462-4 |
| Total cholesterol | 3011163, 3044491, 3007352, 3009966, 3028288, 3028437, 3007070, 3027114, 4008265 | 9830-1, 43396-1, 2091-7, 18262-6, 13457-7, 2089-1, 2085-9, 2093-3, 121868005 |
| Body mass index (BMI) | 40762636 | 59574-4 |
| Weight | 3013762 | 3141-9 |
| Height | 3036277 | 8302-2 |
